# Frequency enrichment of coding variants in a French-Canadian founder population and its implication for inflammatory bowel diseases

**DOI:** 10.1101/2025.07.11.25331388

**Authors:** Claude Bhérer, Jean-Christophe Grenier, Justin Pelletier, Gabrielle Boucher, Genevieve Gagnon, Philippe Goyette, Dariel Ashton-Beaucage, Christine Stevens, Robert Battat, Alain Bitton, Philippe M Campeau, Catherine Laprise, NIDDK IBD Genetics Consortium, Quebec IBD Genetics Consortium, iGenoMed Consortium, Hailiang Huang, Mark Daly, Daniel Taliun, Julie G Hussin, Vincent Mooser, John D Rioux

**Affiliations:** - Department of Human Genetics, Faculty of Medicine and Health Sciences, McGill University, Montréal, Québec, Canada; - Canada Excellence Research Chair Program in Genomic Medicine and Victor Philip Dahdaleh Institute of Genomic Medicine, McGill University, Montréal, Québec, Canada; - Institut de Cardiologie de Montréal, Université de Montréal, Montréal, Québec, Canada; - Medical and Population Genetics, Broad Institute, Cambridge, MA, United States of America; - Department of Gastroenterology, University of Montreal Hospital Center, Montréal, Québec, Canada; - Department of Medicine, McGill University, Montréal, Québec, Canada; - Department of Pediatrics, University of Montreal, Montréal, Québec, Canada; - Département des sciences fondamentales, Centre intersectoriel en santé durable, Université du Québec à Chicoutimi, Chicoutimi, Québec, Canada; - Analytical and Translational Genetics Unit, Massachusetts General Hospital, Boston, MA, United States of America; - Institute for Molecular Medicine Finland (FIMM), University of Helsinki, Helsinki, Finland; - Department of Medicine, Faculté de Médecine, Université de Montréal, Montréal, Québec, Canada; - Mila – Quebec AI Institute, Montréal, Québec, Canada

**Keywords:** founder effect, exomes, genetic association, Mendelian diseases, inflammatory bowel diseases, French Canadian

## Abstract

The genetic features of founder populations with recent bottlenecks, causing some deleterious variants to rise to higher frequencies, can enhance the power of rare variant association studies. French Canadians from Quebec represent a recent founder population with a particular disease heritage comprising more than 30 prevalent Mendelian conditions. Here, we characterize coding variation in this founder population using exome sequencing data from 2,820 French-Canadian participants - patients with inflammatory bowel diseases (IBD), parents and controls from the Quebec IBD cohort. We find that 18% of rare coding variants are 10-100 times more frequent than in non-Finnish Europeans (NFE). A total of 4,133 missense and loss-of-function variants were significantly enriched with a median 28-fold enrichment, revealing the potential for genotype-phenotype associations in this population. We describe significantly enriched pathogenic variants, including those known to account for the increased prevalence of rare diseases in FC compared to other European descent populations, such as Agenesis of corpus callosum and peripheral neuropathy (*SLC12A6*) and Leigh Syndrome French Canadian type (*LRPPRC*). Finally, we investigate whether rare protein-coding variants, enriched in French Canadians by the founder effect, contribute to the risk of IBD using trio and case/control cohorts. In addition to replicating associations in *NOD2* and *IL23R,* we identified new candidate association signals, including enriched variants in *SLC35E3*, and *ARSA.* Our findings show that, even in well-characterized founder populations like the French Canadians, there remains untapped potential for genetic discovery, revealing both rare and complex disease risk factors through enriched coding variation.

## 2 Introduction

Since the early days of human genetics, studies of founder populations have contributed to better understanding Mendelian diseases. The enrichment of rare, deleterious alleles caused by the founder effect may increase the prevalence of certain Mendelian disorders, which in turn facilitates discovery of causal genes and pathogenic variants. Early exemplar studies include linkage disequilibrium (LD)-mapping of diastrophic dysplasia in the Finnish population in 1992^1^, of familial dysautonomia in the Ashkenazi Jewish population in 1993^2^ and the identification of the causal gene and pathogenic variants underlying Leigh Syndrome French Canadian (LSFC) type in this founder population^3^. In contrast, the potential advantages of founder populations for gene discovery have taken considerably more time to be translated in studies of complex traits. Yet, recent studies have demonstrated that novel complex trait association signals often implicate low frequency and rare variants private or enriched in frequency in these populations^4–6^. Here we study the exomes from patients with inflammatory bowel disease (IBD) and from controls (parental and population) in French Canadians, a large population with recent founder effect.

French Canadians from Quebec represent a founder population tracing back to the European colonization that began 400 years ago under French rule^7^. Today, French Canadians include a majority of the 6.5 million French speakers from Quebec^8^. The founder effect in this population is best exemplified by its distinct heritage of Mendelian diseases, with more than 30 monogenic disorders showing increased prevalence in French Canadians compared to other populations^9–11^. The population history, including the initial peopling of Nouvelle-France and the subsequent demographic and spatial expansions, is thought to have elicited serial founder effects that impacted to various degrees the genetic make-up of regional and ethno-cultural subpopulations of Quebec^12–16^. Previous genomic studies have shown that French Canadians are genetically differentiated from continental Europeans, showing increased frequency of some deleterious variants^13–15,17,18^. Yet these studies have been limited to genotyping data, low coverage sequencing or small-scale deep sequencing datasets, which only partially described the rare variant distribution in this population. Consequently, studies of coding variation in larger samples of French Canadians are still needed to better understand the consequences of the founder effect for rare and complex diseases.

To address this, we performed exome sequencing in 3,102 individuals living in the Canadian Province of Quebec. Participants in this study were patients with IBD, parents or controls, included in the Quebec IBD Cohort. IBD, consisting primarily of Crohn’s disease (CD) and ulcerative colitis (UC), are chronic inflammatory diseases of the gastrointestinal tract. We analyzed a subset of 2,323 unrelated French-Canadian individuals to assess the patterns of coding variation in the French-Canadian population of Quebec, and in the entire French-Canadian cohort to evaluate the potential of this founder population’s characteristics for the discovery of susceptibility. We focused on IBD for multiple reasons. First, IBD has been a prototypical complex trait with early discovery of causal genes by positional cloning (e.g. *NOD2*) and genome-wide association (e.g. *IL23R*) approaches^19–21^. Since these early studies, genome-wide association (GWAS) and sequencing studies have led to the identification of over 200 loci, with putative or functionally confirmed causal coding variants identified in multiple genes^22–26^. Second, Canadians in general including French Canadians have among the world’s highest prevalence of IBD, currently estimated to be 825 per 100,000^27^. Third, participants from the French-Canadian population have contributed to prior successful genetic discovery efforts in IBD^22,26,28–30^. In this study, we describe the enrichment of coding variation in the French-Canadian population of Quebec, including pathogenic variants associated with Mendelian diseases (some previously described as founder variants in this population) and in a subset of known and candidate IBD-associated variants. We discuss potential implications of this signature of the founder effect on our understanding of disease burden in this population and on genetic discovery efforts.

## 3 Material and Methods

### 3.1 The Quebec IBD cohort

The study participants were recruited in the province of Quebec, Canada. The Quebec IBD cohort includes participants from three different studies: The IBDGC-Montreal, the iGenoMed-MTT, and the Genome Quebec-GENIZON biobank studies. Supplementary Table 1 describes the number of samples included from each study. **IBDGC-Montreal:** This Montreal-led Genetic Research Center (GRC) is one of the seven GRCs of the NIDDK Inflammatory Bowel Disease Genetics Consortium (IBDGC). Patients and controls (spouses or best friends) for this study were recruited at university-based hospitals throughout the province by gastroenterologists specializing in IBD that formed the Quebec IBD Genetics Consortium. **iGenoMed-MTT:** is an ongoing prospective study of patients with a confirmed diagnosis of IBD that are followed longitudinally and evaluated for their response to molecularly targeted therapies (MTT). Patients have been recruited in Montreal at the Centre hospitalier de l’Université de Montréal (CHUM) and the McGill University Health Centre (MUHC). **Genome Quebec-GENIZON:** Samples were previously collected by Genizon Biosciences Inc. but are now part of a public biobank overseen by Genome Quebec. Participants in this study include patients with Crohn’s and unaffected parents, or unrelated controls^31^. For all cohorts, IBD (Crohn’s disease, ulcerative colitis) was diagnosed according to accepted clinical, endoscopic, radiological and histological findings^31,32^. Control individuals were selected to match the patient’s regional distribution.

### 3.2 Ethics statement

All participants from studies included in the Quebec IBD Cohort provided informed consent. Recruitment protocols and consent forms were approved by institutional review boards at each participating institution. Study protocols were approved by the Montreal Heart Institute Institutional Ethics Committee « Comité d’éthique de la recherche et du développement des nouvelles technologies » (IBD Genetics 2005-23 (05-813) and MTT: MP-02-2017-7170, 2017-2202) and the Research Ethics Office (IRB) of the Faculty of Medicine and Health Sciences at McGill University, Canada (IRB Number A01-M04-21A).

### 3.3 Exome sequencing and production of initial variant call set

Exome sequencing was performed at the Broad Institute, Cambridge MA, USA, as previously described^26^. Sample library preparation was carried out using Illumina Nextera followed by hybrid capture (Illumina Rapid Capture Enrichment Nextera, 37Mb target, and Twist Custom Capture, 37 Mb target). Sequencing was performed on HiSeqX instruments to 150-bp paired reads. Sequencing was performed at a median depth of 85% targeted bases at > 20X. Sequencing reads were mapped by BWA-MEM to the GRCh38 human reference genome using a ‘functional equivalence’ pipeline. Mapped reads were then marked for PCR and optical duplicates, and base quality scores were recalibrated. They were then converted to CRAM using Picard 2.16.0-SNAPSHOT and Genome Analysis Toolkit (GATK) 4.0.11.0^33^. The CRAMs were then further compressed to gVCFs using ref-blocking. These were used for joint variant calling, processing separately for samples sequenced using Nextera and Twist captures, in order to create an initial set of raw single-nucleotide variants (SNVs) and short insertions/deletions (InDels) calls. Variant call accuracy was estimated using the Genome Analysis Toolkit variant quality score recalibration (VQSR) approach^34^. Quality control of this initial call set included exclusion of samples with ≥ 10% contamination, and of those with less than 40% of targets at 20X coverage.

### 3.4 Exome sequencing data quality control and variant filtering

Starting with the initial call-set described above (n=3,102 samples), we performed individual-level and variant-level filtering. We excluded samples with contamination rate > 2% (n=32) and chimeric rate > 1.5% (n=1) (Supplementary Table 2). Multi-allelic sites were split into biallelic variants and InDels were normalized using bcftools v1.9^35^. Using a set of LD-pruned variants with minor allele frequency (MAF) >0.01, allowing for 5% missingness, duplicated samples were identified and for each duplicated pair, one sample was kept using PRIMUS v1.9.0^36^. A total of 28 duplicated samples were removed (Supplementary Table 2). Using the same set of variants, we inferred genetic sex using exome sequencing data using PLINK v.1.9^37^ and excluded samples with discordant inferred genetic sex and self-reported sex at birth (n=26) (Supplementary Figure 1). Variant-level filtering was performed separately for the autosomes, pseudoautosomal regions and sex chromosomes. For autosomes and pseudoautosomal regions, the heterozygous calls were filtered according to their allele balance, retaining variants with allele balance between 30% and 70%. Genotypes with a depth less than 10 were set as missing, Mendelian errors were identified with the trio information we had in the cohort using PLINK v1.9^37^ and set to missing in those samples only (option -me 1 1). Variants with more than 5% of missing rate were then removed, those below a Hardy-Weinberg (HWE) threshold of p < 1 x 10-6 (--hwe 0.000001 midp) and positions outside the intersection of the two target kits used +/-50bp were also removed. The non-pseudoautosomal region (non-PAR) of the X chromosome and the Y chromosome were filtered separately. The heterozygous calls for the males were filtered out and only calls for males were kept for the Y chromosome. The centromeres were removed and calls with coverage of less than 10 were removed for females and a threshold of 5 was considered for the males. For the non-PAR region of X chromosome using both males and females, Mendelian errors were assessed, but none were identified, positions with missing rate higher than 5% were removed, the same HWE filter as for the autosomes was applied and only the positions matching the capture kit (+/- 50bp around the target regions) were kept. For the Y chromosome, the same filters were applied except for Mendelian errors and HWE filters. Finally, the genotypes on the mitochondrial chromosome were filtered for a minimum depth coverage of 5, maximum missing rate of 5% and positions matching the capture kit (+/- 50bp around the target regions).

### 3.5 Reference datasets

We used two reference datasets: the 1000 Genomes Project and the Quebec regional reference panel (ERRQ)^13,38^. For the 1000 Genomes reference dataset, we used the high coverage ( ≥ 30x) whole genome sequencing (WGS) data sequenced at the New York Genome Centre^39^. Specifically, we used a subset of 2,504 unrelated individuals from 26 globally diverse populations (Supplementary Table 3). Variants with a MAF of less than 1% were removed, and only variants with more than 99% of genotypes (< 1% missingness) were retained for principal component analyses (below). As reference set for French Canadians from Quebec, we used the ERRQ reference panel, which comprise individuals self-reported with 3 or 4 grandparents from 10 different regional and ethno-cultural origins (Supplementary Table 4)^13^, including 232 unrelated individuals from the Saguenay−Lac-St-Jean asthma familial collection^38^. After quality control filtering, we kept 741 of the 745 genotyped ERRQ samples from the NeuroX genotyping array and 309 of the 313 samples genotyped on an Omni array, for a total of 620,089 variants.

### 3.6 Population structure and relatedness

We performed principal component analysis (PCA) on the 1000 Genomes Project’ samples using FlashPCA2^40^. We used LD-pruned variants (sliding windows of 50Kb, moving by 5 variants steps with a 0.5 r^2^ threshold) with missingness < 0.01 and MAF > 0.01. Samples from the Quebec IBD cohort were projected on the 1000 Genomes PCA, using a set of variants in common between the two datasets. We used a k-nearest neighbors approach applied on PCA projections of the Quebec IBD cohort samples on the 1000 Genomes reference PCA to identify individuals most genetically similar to European/non-European samples in the 1000 Genomes. Specifically, for each individual in the Quebec IBD cohort, we computed the Euclidean distances with the 1000G reference samples using the top 20 PCs and identified the k=10 nearest reference samples in this PCA space. To identify Quebec IBD cohort individuals most genetically similar to Europeans, we used a threshold 8 out of 10 nearest reference samples from the 1000 Genomes non-Finnish European samples. Remaining individuals, with 3-10 nearest reference individuals from non-European 1000 Genomes samples, were defined as having ancestries from non-Europeans (the non-EUR subset). Likewise, to identify individuals most genetically similar to French-Canadian reference samples, we created a reference set combining samples from the ERRQ (n=499) and non-Finnish Europeans from the 1000 Genomes (n=404). For this combined ERRQ and 1000 Genomes European reference set, we created a set of LD-pruned variants with MAF> 1% from the variants in common between the ERRQ genotyping array data and the 1000 Genomes. We computed 20 PCs on this combined ERRQ and 1000 Genomes reference set and projected the Quebec IBD cohort samples inferred as genetically closer to Europeans. Individuals more genetically similar to 1000 Genomes European samples were defined as the European non-FC subset (EUR-non-FC), based on k-nearest neighbor approach using a threshold of 8 out of 10 nearest individuals. Remaining individuals, with a larger number of nearest reference samples from the ERRQ panel were defined as more genetically similar to French Canadians and included in the FC subset. To identify individuals unrelated up to the third degree we used PRIMUS v1.9.0^36^, which algorithm identifies the subset of unrelated individuals that maximizes sample size.

### 3.7 Variant annotation

Variants were annotated using Variant Effect Predictor (VEP) (version 109)^41^. Loss-of-function predictions were assessed using the LOFTEE plugin^42^. LOFTEE considers stop-gained, splice-disrupting, and frameshift variants, and filters out many known false-positive modes, such as variants near the end of transcripts and in non-canonical splice sites. ClinVar database (version 202209) was used to annotate variants’ clinical significance^43^.

### 3.8 Allele frequency enrichment analyses

For each variant, we computed the MAF enrichment as the MAF ratio between a given subset of French-Canadian exomes (UNRFC and FC controls) and gnomAD NFE exomes v2.1. To test for MAF enrichment in the French Canadian subsets, one-sided Fisher exact tests were computed using the minor and major allele counts in a given subset of French-Canadian exomes and gnomAD NFE exomes. We also computed biased corrected odds ratios, adding 0.5 to minor and major allele counts following Rivas et al.^44^, to obtain enrichment values for variants with zero allele counts in gnomAD NFE.

To evaluate regional enrichment of known pathogenic variants, we first established cutoff values on PC1 and PC2 from the ERRQ reference set for the Saguenay−Lac-Saint-Jean (SAG) and North Shore (NS), and Gaspesia Acadian (ACA) regions. The SAG region was identified as individuals with 0.025 ≤ PC1, while the NS region was defined as individuals with PC1 ≤ −0.025 and 0.01 ≤ PC2 (**Figure 1**). Individuals were assigned to a specific region based on their PCA projection coordinates within the predefined ERRQ reference space. For each pathogenic variant, we constructed contingency tables comparing the number of carriers within the region to those outside it. We then applied chi-squared tests to assess whether the variant was significantly enriched in that region.

**Figure 1.**
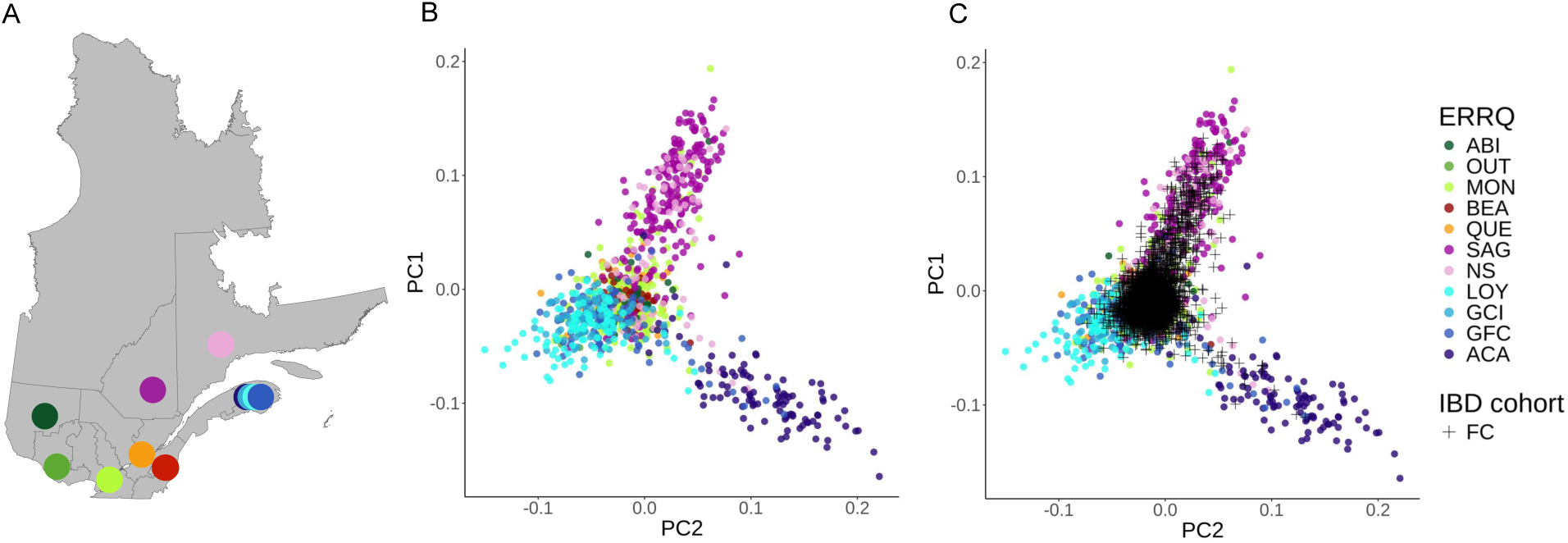
**Fine-scale structure of participants in the FC subset of the Quebec IBD cohort** (A) Geographical location of the 11 regional or ethno-cultural groups sampled in the ERRQ Reference Panel in the Province of Quebec (Canada). (B) PCA of the ERRQ Reference Panel. PC1 (1.97% of total variance explained) is shown against PC2 (1.45% of total variance explained) (C) PCA Projection of the FC subset. Abbreviations: ABI: Abitibi-Témiscamingue; OUT: Outaouais; MON: Montreal; BEA: Beauce; QUE: Quebec City; SAG: Saguenay−Lac-Saint-Jean; NS: North-Shore; LOY: Gaspesia Loyalist; GCI: Gaspesia Channel Islanders; GFC: Gaspesia French Canadians; ACA: Gaspesia Acadians.

### 3.9 Birth prevalence estimation

Birth prevalence was estimated using the Quebec IBD cohort exome data and clinical data for one example disease, namely Cartilage-Hair Hypoplasia (CHH), an autosomal recessive disorder caused by pathogenic variants in the *RMRP* gene. First, to estimate prevalence from exome data, we extracted all variants in the *RMRP* gene classified as pathogenic or likely pathogenic (PLP), or with conflicting interpretations of pathogenicity according to ClinVar. We then performed manual curation of these variants, excluding variants with conflicting interpretations without other PLP classifications. Curated variants were retained to compute the cumulative frequency of variants and compute the disease birth prevalence, assuming Hardy-Weinberg equilibrium and complete prevalence of PLP variants.

Second, we estimated the birth prevalence of CHH using clinical data. A total of five patients diagnosed with CHH at CHU Sainte-Justine between 2013 and 2023 were identified. The study was conducted in accordance with the Declaration of Helsinki and approved by the Ethics Committee of CHU Sainte-Justine (protocol code 015-853:4072 and date of approval 26 May 2015). Information was shared according to the regulation in place at the CHU Sainte-Justine. Birth prevalence from these clinical data was obtained by dividing the number of observed cases, by the number of births reported in Quebec^45^. In order to get an estimate for French-Canadians specifically, we assume that the number of births represents 80% of total births, which is approximately the proportion of the French speaking population in the province^8^. We assumed the number of births to be 40% of French-Canadian live births reported in Quebec during the 10-year period, since this proportion reflects the population coverage by this tertiary care center^46^.

### 3.10 Association testing

Single variant association test was conducted using the Sequence Kernel Association Test (SKAT) implemented in the SAIGE (Scalable and Accurate Implementation of Generalized mixed model) software^47^ (version 1.1.6.3). We performed SKAT single variant association tests in cases and controls that are not part of the trios from the FC subset inferred most genetically similar to French Canadians from the ERRQ reference panel (n=1,776) (Supplementary Table 5). Tests were performed on a total of 192,087 variants that passed quality control procedures (above) and MAF > 0.001. To control for sub-population structure and potential confounding effects, we performed PCA using FlashPCA2 software^40^ and a set of well-genotyped common LD-pruned variants (passing all above QC filters, with MAF > 0.01, call rate > 99%), excluding regions with known long-range linkage disequilibrium (LD). PCA was performed on individuals unrelated up to the third degree and related individuals were projected on these PCs. SKAT single variant association tests were performed for IBD and CD only using the same set of non-IBD controls, controlling for sex and top 4 PCs.

A Transmission Disequilibrium Test (TDT) using Plink (version 1.9) was conducted on French-Canadian mother-father-affected child trios (n=954). The test uses deviations from Mendelian inheritance patterns to identify potential genetic associations^48^. The TDT test is a test of linkage in the presence of association between the trait and the genotype.

Results from the TDT and single variant association were combined using an inverse-variance approach. For the TDT, effect size was computed as the transmitted to untransmitted ratio, after correction for continuity (adding 0.5 to both), while the variance estimate was derived from the chi-squared statistic and effect size.

Finally, a set-based association test was conducted using SAIGE-GENE+ v1.1.6.3^49^ using the same set of FC individuals as the single-variant association test (n=1,776). The annotation masks for each set were: “--annotation_in_groupTest=“LoF:non-synonymous,synonymous”. We ran set-based association tests with MAF filters of 0.001, 0.01, and 0.05.

## 4 Results

### 4.1 French-Canadian exomes in the Quebec IBD cohort

To gain knowledge of coding variation patterns in the French-Canadian population and its implications for disease association, we analyzed exome sequencing data generated for 3,102 participants from the Quebec IBD cohort. Sequencing was conducted at the Broad Institute as part of an international collaborative sequencing study being performed under the umbrella of the International IBD Genetic Consortium^26^. After quality control (see Material and Methods, Supplementary Tables 1-2), we obtained a call set of 3,015 exomes comprising 848,032 small variants, including 804,034 SNVs and 43,998 InDels (Supplementary Tables 6-7). Among these, 347,354 were singletons (41%), and 52,994 (6.25%) were novel variants, i.e. not found in dbSNP.

To assess genetic similarity to reference populations, we projected exomes from participants in the Quebec IBD cohort onto a PCA space defined by the 1000 Genomes WGS reference dataset (see Material and Methods). In the 1000 Genomes PCA space built with 2,504 participants across 26 worldwide populations, most IBD cohort participants clustered near European reference samples (Supplementary Figure 2, Supplementary Table 3). Using a k-nearest neighbors approach applied to this PCA projection, we identified 2,947 individuals most genetically similar to European samples from the 1000 Genomes Project, and 68 individuals with non-European genetic ancestries (the non-EUR subset) (Supplementary Table 7), reflecting the multi-ancestry sampling of the Quebec IBD cohort. To further subset individuals most genetically similar to the French-Canadian founder population, we projected the 2,947 samples onto a PCA defined by a combined set of NFE from the 1000 Genomes Project and the ERRQ reference panel, a genotype reference dataset ascertained for geographic and ethno-cultural origins in Quebec^13,38^ (Supplementary Figure 3, Supplementary Table 4). As expected, given that participants were recruited in Quebec, most individuals clustered with the ERRQ samples (Supplementary Figure 3). Using a k-nearest neighbors’ approach, we identified 2,820 individuals, defined as the FC subset, most genetically similar to the French-Canadian reference individuals from the ERRQ than to the 1000 Genomes Project Europeans. The remaining 127 individuals formed the EUR-non-FC subset. The FC subset comprised 2,820 exomes containing 653,574 small variants, including 623,119 SNVs and 30,455 InDels (Supplementary Table 7) and was used for association testing (see below). Finally, from the 2,820 individuals of French-Canadian genetic ancestry, we generated a subset of 2,323 individuals unrelated up to the third degree, defined as the unrelated French-Canadian (UNRFC) subset.

To characterize patterns of genetic variation within the French-Canadian population, we used the FC and UNRFC subsets. French-Canadian exomes in the FC subset show a mean number of 44,053 SNVs (539.6 s.d. or 95% CI) and 1,969 InDels (49.6 s.d. or 95% CI) (Supplementary Table 9). These numbers of variants are significantly lower than the EUR-non-FCs (mean= 44,462 SNVs, pairwise Wilcoxon test W = 283,693 and p-value = 36.9 x 10^-29^ for SNVs and InDels) and non-EUR subsets (mean= 46,456 SNVs, pairwise Wilcoxon test W = 1,5183.5 and p-value = 1.6 x 10^-32^ for SNVs and InDels) (Supplementary Figure 4, Supplementary Table 9), in line with the hypothesis that the French-Canadian founder effect led to a loss of genetic variation. Our PCA analyses of the FC subset along with ERRQ samples recapitulate known fine-scale genetic structure in the French-Canadian population (**Figure 1**, Supplementary Figures 5-7). The primary axis of variation corresponds to a west-to-east gradient, with individuals from urban centers of Montreal and Quebec City clustering separately from eastern regional populations. The two most differentiated regional groups comprise the Northeast (represented by the Saguenay−Lac-Saint-Jean and North Shore samples in the ERRQ reference panel), and the Acadians from Gaspesia, which are known to display the strongest signatures of the founder effect in Quebec^12–14^. Hence, the Quebec IBD cohort includes a majority of participants clustering with reference samples from urban centers and also includes individuals clustering with the Northeast and Gaspesia Acadians, enabling more fine-scale analyses.

### 4.2 Enrichment of coding variation in Mendelian disorders genes in French Canadians

To characterize the French-Canadian coding variation, we compared the allele frequencies of coding variants observed in the UNRFC (N= 2,323) to those from NFE in gnomAD exomes (Material and Methods). As expected, due to the predominant European ancestry of French Canadians^12^, most variants observed in French-Canadian exomes are in common with NFEs specifically 86.5% of 398,626 coding variants (Supplementary Table 10). As such, only a small fraction of variants observed in the UNRFC exomes are not in gnomAD NFE exomes, with most of these variants (82.3%) being singleton variants in the UNRFC. Likewise, 95.5% observed in the UNRFC subset are found in dbSNP, and 90.8% of variants not in dbSNP, or novel variants, were singletons. Novel variants were equivalently distributed across the different functional categories (Supplementary Table 10). To identify alleles with evidence of enrichment within the French-Canadian population, we compared the MAF of these variants between the UNRFC and NFE. We observed that over 20% of protein-coding variants had at least 10-fold frequency enrichment in the UNRFC dataset (**Figure 2A**). This enrichment was observed for all types of coding variants albeit with a greater fraction of enriched missense variants (31.1%) and putative LoF variants (45.5%) as compared to synonymous variants (24.2%) (**Figure 2B-D**). As expected, enrichment in the UNRFC subset relative to NFE is stronger for ultra-rare (allele count (AC) < 4) and rare variants (0.001<MAF<0.01) compared to intermediate frequency (0.01<MAF<0.05) and common variants (MAF>0.05) (Supplementary Figure 8).

**Figure 2.**
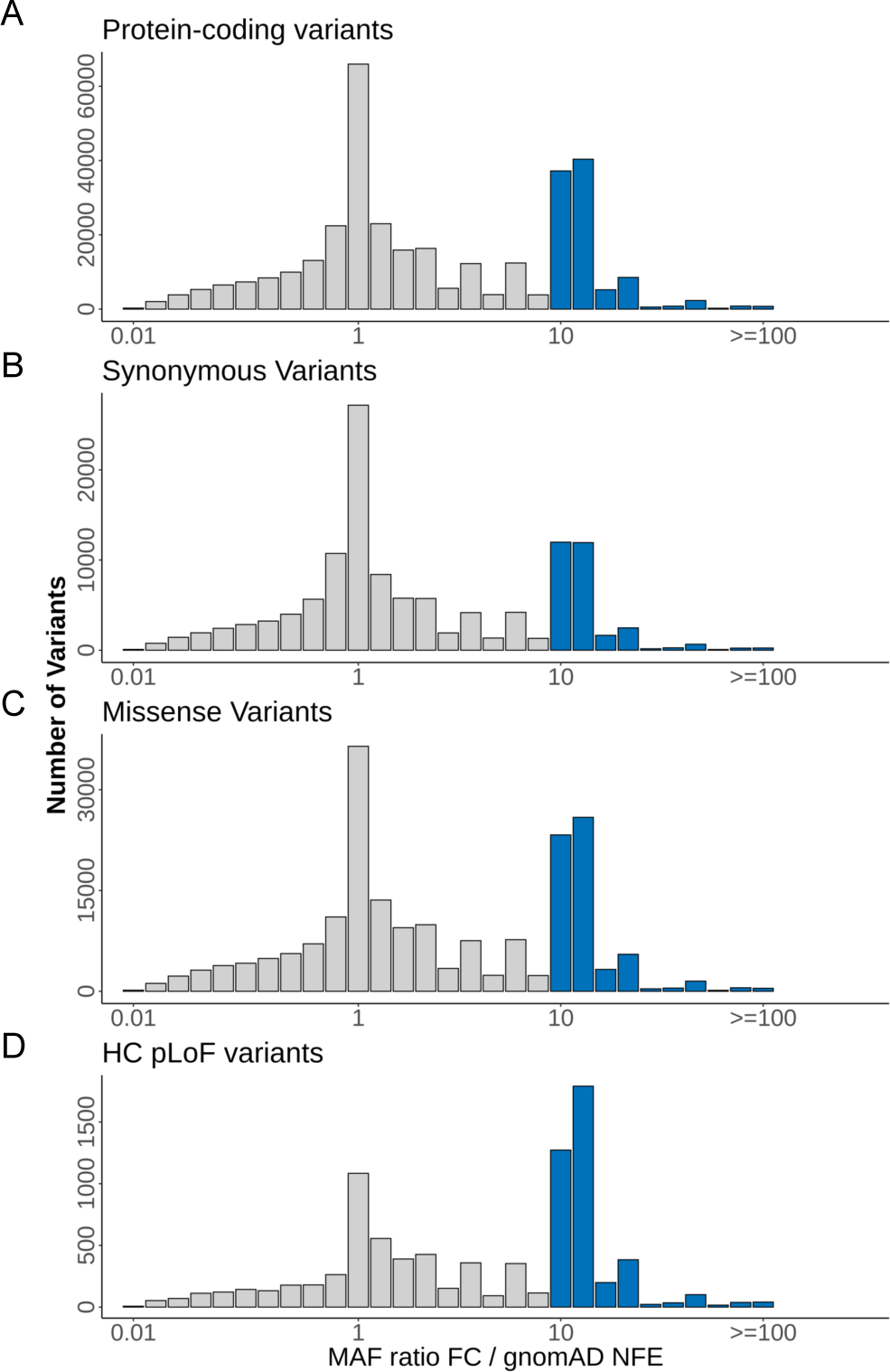
**Frequency enrichment of protein-coding variants in French-Canadian exomes**. Histograms show the distribution of variants as a function of their MAF ratio in the UNRFC sample relative to gnomAD NFE exomes. Variants with MAF ratio equal or above 10 in the UNRFC are highlighted in blue. Frequency enrichment histograms are shown for (A) all protein-coding variants, (B) synonymous variants, (C) missense variants, (D) high-confidence (HC) pLoF variants.

As much of the genetic characterization of the French-Canadian founder population to date has focused on Mendelian disorders, we compiled a list of 72 pathogenic variants, implicated in 31 diseases, that were previously described as candidate founder variants^9–11^ (Supplementary Table 11). Among those, we identified 37 pathogenic variants (implicated in 21 diseases) in exome data from the UNRFC subset (**Figure 3A**). Significant enrichment is observed for 19 of these 37 previously described founder variants in the UNRFC subset relative to NFE (one-sided Fisher exact p-value < 0.05), and 84% (16 of 19) of these show a strong enrichment (equal or above 10-fold), reflecting the high prevalence of these pathogenic variants in the population (**Figure 3A**; Supplementary Table 11). Some of the most notable examples include the pathogenic variant in *SLC12A6*, the causal gene for agenesis of corpus callosum with peripheral neuronopathy (ACCPN) (c.2436+1delG) with 76-fold enrichment (one-sided Fisher exact test p-value = 9.19 x 10^-37^), c.8844del pathogenic variant in *SACS* the causal gene for Spastic Ataxia of Charlevoix-Saguenay (ARSACS) (26-fold enrichment, one-sided Fisher exact test p-value =5.53 x 10^-15^), c.1061C>T variant in *LRPPRC* implicated in LSFC (26-fold enrichment, one-sided Fisher exact test p-value = 1.73 x 10^-12^) and c.1062+5G>A in *FAH* implicated in tyrosinemia type 1 (4-fold enrichment, one-sided Fisher exact test p-value =1.49 x 10^-4^) (Supplementary Table 11). These four pathogenic variants are implicated in canonical French-Canadian founder effect diseases and are part of a prenatal carrier screening program targeting a specific regional population of Quebec^50^. As such, our results confirm a regional enrichment of carriers of these four pathogenic variants in the North-Eastern regions of Quebec (**Figure 3B**, Supplementary Figure 9). Specifically, carriers of these pathogenic variants cluster significantly with the Saguenay−Lac-Saint-Jean and North Shore regional samples in the PCA projections based on the ERRQ reference panel (chi-square p-value of carriers in North-East vs outside < 0.05).

**Figure 3.**
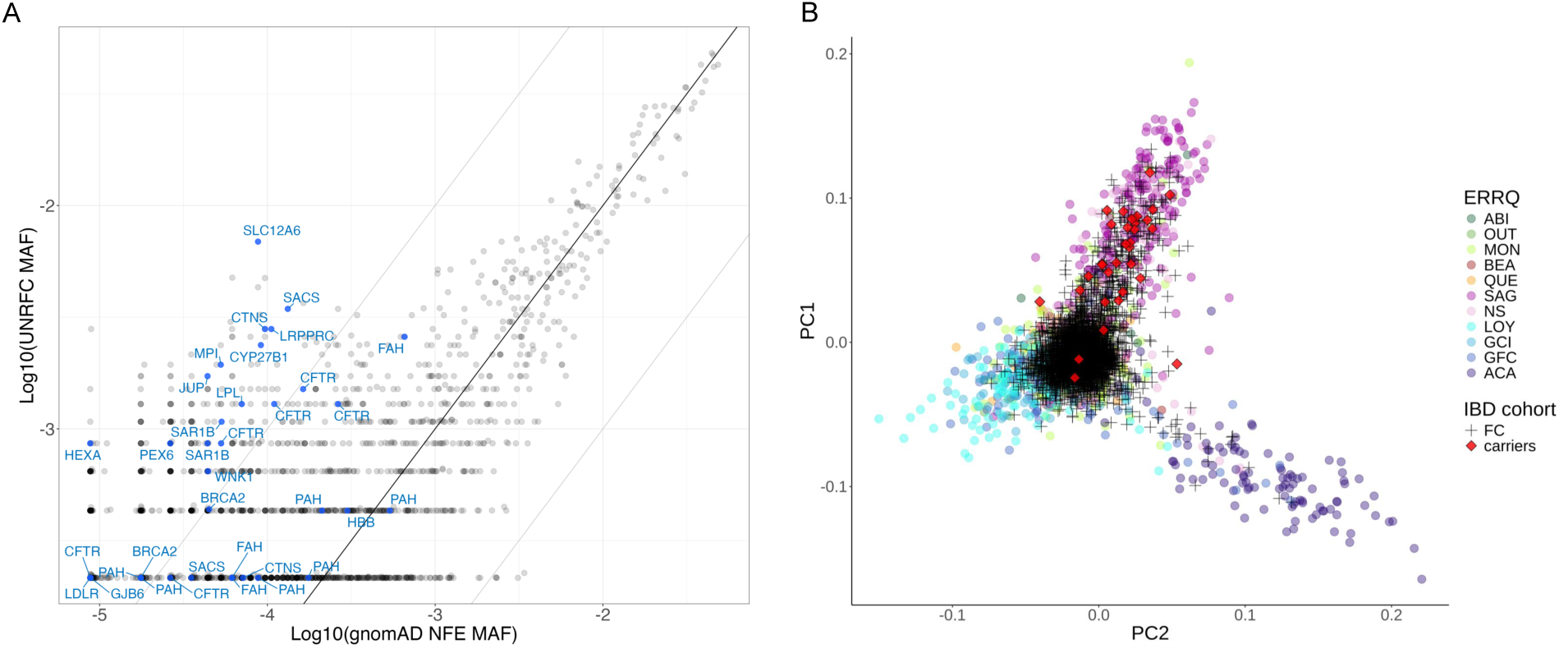
**Enrichment of pathogenic variants in French-Canadian exomes**. (A) Scatterplot of MAF of ClinVar PLP variants (gray) and previously described French-Canadian founder variants (PLP) in the UNRFC subset relative to gnomAD NFE (B) PCA projection of the UNRFC subset in the ERRQ PCA space, highlighting in red the carriers of *SLC12A6* c.2436+1delG pathogenic variant implicated in ACCPN.

To expand beyond this analysis, we assessed the enrichment of all variants with ClinVar interpretation as ‘pathogenic’ or ‘likely pathogenic’ (PLP) in the UNRFC compared to NFE (**Figure 3**, Supplementary Table 12), excluding all previously associated IBD genes^22,51^, in order to assess pathogenic variant enrichment independent of IBD-specific signals. Among those 15.7% of 2,316 PLP (n=363 PLP variants) had at least 10-fold enrichment, suggesting that there is an important number of clinically relevant variants enriched in the French-Canadian population of Quebec, beyond those which have been already described. The top 10 enriched PLP variants showed an enrichment > 61-fold in the UNRFC subset. Amongst the enriched ClinVar PLP, we find well-established disease-causing variants with 2 stars interpretations that were not described in a Quebec context yet, such as *LAMA3* variant c.9511+1G>A implicated in Junctional epidermolysis bullosa gravis of Herlitz (biased-corrected OR=73.3, one-sided Fisher exact test p-value=1.69 x 10^-5^), and nonsense variant p.Arg124Ter in *FRAS1* implicated in Fraser Syndrome 1 (biased-corrected OR=17.4, one-sided Fisher exact test p-value=2.40 x 10^-6^). Our enrichment results hence point these as candidate founder variants. Amongst enriched PLP variants, however, we find several variants with conflicting interpretations, showing that a systematic curation would be required to get an accurate picture of enriched pathogenic variants in the French-Canadian population. While we detected significant enrichment of the known founder effect variants and many of the ClinVar variants in the entire UNRFC subset, not surprisingly we observe regional differences in carrier frequencies (Supplementary Figure 9). For example, carriers of the known causal variants in the *LRPPRC*, *SACS*, and *SLC12A6* genes cluster almost exclusively with the SAG reference samples, *USH1C* carriers cluster with Acadians from Gaspesia and “central Quebec” (QC, MTL), and *FRAS1* almost exclusively in “central Quebec” (QC, MTL).

Looking at a less stringent enrichment threshold, there are 376 PLP variants with 4-10X fold enrichment in the UNRFC subset relative to NFE. While most of these are singleton variants (55.05%), 28.99% are shared by 2-4 carriers in the UNRFC, and 15.96% by 5 carriers or more, again consistent with a much larger number of founder variants yet to be described as so in the French-Canadian population. An notable example is *RMRP*, a gene known to be involved in cartilage-hair hypoplasia (CHH), an autosomal recessive congenital disorder which is found across Europe but shows increased frequency in the Finnish and Old Order Amish populations^52–54^. In the UNRFC subset, we identified 15 carriers of 4 pathogenic variants in *RMRP*, including 3 singletons (Supplementary Table 13). Only one variant in *RMRP* (n.70A>G) is significantly enriched in the French-Canadian exomes (AF=0.00258, 4.26-fold enrichment, one-sided Fisher exact p-value = 1.85 x 10^-4^). The enriched variant n.70A>G represents 80% (12/15) of carriers of pathogenic variants in *RMRP* in the UNRFC subset, corresponding to a carrier frequency of 1:194. By using the cumulative carrier frequency over the 4 pathogenic variants in UNRFC and assuming Hardy-Weinberg equilibrium, the birth prevalence of CHH is estimated to be 1.04 per 100,000 (95% CI [0.22, 5.46]) or 1 in 95,935 births in the French-Canadian population. To assess if this estimate is consistent with French-Canadian affected individuals’ data within the Quebec population, we investigated CHH probands identified at the CHU Sainte-Justine tertiary care center, covering 40% of Quebec’s pediatric population^46^. We identified 5 affected individuals from 4 families diagnosed with CHH between 2013 and 2023 in this center, in a database of individuals with congenital disorders. The n.70A>G variant accounted for 75% (6/8) of the disease-causing variants in *RMRP* from parents with self-reported French-Canadian ancestry (Supplementary Table 14). From the number of probands with CHH identified in this tertiary care center, we estimated a birth prevalence of 1.34 per 100,000 births or 1 in 74,247 births, which is consistent with our estimate based on exome data.

### 4.3. Complex trait discovery: IBD association analyses

To evaluate the potential of the French-Canadian founder population for the study of complex traits we have focused on IBD. The high incidence and prevalence of CD, UC, and IBD in the province of Quebec^55–57^ offer an opportunity to investigate how the founder effect has impacted coding variants associated with IBD risk. To address this question, we evaluated the genetic association in the FC subset of the Quebec IBD cohort of previously validated causal variants in the coding sequence within known IBD GWAS loci. Specifically, we examined our dataset for the presence of 25 variants within 14 causal genes, with all but one gene (*RNF186*) being associated with CD and/or IBD in the current dataset (**Table 1**, Supplementary Data 2). Among these, 24 variants were detected in our cohort and one was removed from the analyses (c.IVS17+5A>G in gene *CUL2*) during quality control steps due to excess heterozygosity. Next, we performed a TDT analysis in the 318 complete mother-father-affected child (all having CD) trios, as well as a case-control analysis in the independent set of 783 CD, 249 UC and 761 unrelated controls, followed by a combined analysis of these results. The most significant associations with the greatest genetic effect were found in the *NOD2* and *IL23R* genes (Table 1 and Supplementary Table 15). Notably, four of the six of the known variants in *NOD2* that were detected in our cohort were significantly enriched in French Canadians (1.5- to 2.4-fold enrichment in the FC controls subset relative to gnomAD NFEs, one-sided Fisher exact test p-value < 0.05), and five had significant association to CD/IBD and conferred increased risk. All three known causal variants in the *IL23R* gene were detected in this cohort. Two of these were significantly associated with a protective effect in IBD, and one was found to be significantly enriched (IL23R*V362I). We also observed that one of the variants in the *IFIH1* was significantly enriched (2.73-fold) in French Canadians and was associated with IBD.

**Table 1.** Summary of top single point association test results.

| Gene Symbol | Variant | Impact of variant | MAF |  | Enrichment |  | Combined IBD |  | Combined CD |  |
| --- | --- | --- | --- | --- | --- | --- | --- | --- | --- | --- |
|  |  |  | NFE | FC Ctrl | OR | p-value | BETA | p-value | BETA | p-value |
| **IL23R** | 1:67240275:G:A | p.Arg381Gln | 0.059 | 0.061 | 0.96 | 6.60E-01 | -0.68 | 5.03E-06 | -0.70 | 4.58E-06 |
| **IL23R** | 1:67240217:G:A | p.Val362Ile | 0.017 | 0.024 | 1.5 | 1.73E-02 | -0.79 | 4.25E-04 | -0.71 | 1.93E-03 |
| **NOD2** | 16:50712015:C:T | p.Arg702Trp | 0.043 | 0.084 | 2.03 | 3.81E-11 | 0.54 | 6.53E-08 | 0.65 | 1.03E-10 |
| **NOD2** | 16:50712288:G:A | p.Val793Met | 0.002 | 0.001 | 0.72 | 7.63E-01 | 1.13 | 5.71E-02 | 1.28 | 3.36E-02 |
| **NOD2** | 16:50729867:G:GC | fs1007insC | 0.023 | 0.035 | 1.58 | 2.02E-03 | 0.97 | 2.77E-14 | 1.11 | 4.32E-19 |
| **NOD2** | 16:50722629:G:C | p.Gly908Arg | 0.014 | 0.021 | 1.5 | 2.65E-02 | 0.62 | 1.21E-03 | 0.75 | 7.03E-05 |
| **LTA** | 6:31572779:T:C | p.Cys13Arg | 0.267 | 0.246 | 0.90 | 9.52E-01 | 0.34 | 4.51E-06 | 0.37 | 8.16E-07 |
| **SLC35E3** | 12:68746559:G:C | p.Cys61Ser | 0.023 | 0.040 | 1.67 | 4.85E-04 | -0.69 | 6.94E-05 | -0.64 | 3.58E-04 |
| **ARSA** | 22:50625182:C:T | p.Arg498His | 0.040 | 0.078 | 2.03 | 2.47E-10 | -0.45 | 2.56E-04 | -0.40 | 1.49E-03 |

Given this evidence of association and enrichment for variants from a subset of known IBD causal genes, we next evaluated the evidence in the exome sequencing data for potentially novel loci in French Canadians. In this exploratory discovery phase, we examined the combined analysis results for variants with an association p-value <1 x 10^−3^. Thirty-one variants met this criterion (Supplementary Table 16). These variants were in 29 different genes, with ten of these being clustered in two genomic regions. The two most significant associations were for the p.Cys13Arg variant in the *LTA* gene (P_IBD_=4.19 x 10^-6^; P_CD_= 9.50 x 10^-7^), and the p.Cys61Ser variant in the *SLC35E3* gene (P_IBD_=5.84 x 10^-5^; P_CD_= 3.15 x 10^-4^). A conditional logistic regression analysis performed on the *LTA* variant identified a second independent signal in the region, centered mostly in the region containing the class II major histocompatibility genes. While there was no enrichment detected for the variant in the Lymphotoxin alpha (*LTA*) gene, there was significant enrichment detected for the variant in the Solute Carrier Family 35 Member E3 (*SLC35E3*) gene (fold-enrichment=1.67, one-sided Fisher exact test p-value=4.85 x 10^−4^). Moreover, in the other regions we found significant enrichment for multiple other variants in this set of suggestively associated variants (P<10^−03^) with the top one being the p.Arg498His variant in the arylsulfatase A (*ARSA*) gene (enrichment: OR=2.03, one-sided Fisher Exact test p-value=2.47 x 10^−10^). In terms of biological functions related to these genes, the *LTA* gene encodes lymphotoxin A, a cytokine secreted by multiple immune cells, with the associated coding variant (p.Cys13Arg) located in the signal sequence of this gene, potentially impacting on its ability to be secreted (see Table 1). It is believed that the transmembrane golgi protein encoded by *SLC35A3*, a member of the solute carrier family, has UDP-GlcNAc transporting activity and contributes to the fine tuning of inflammatory processes (Table 1)^58^. *ARSA* is involved in sphingomyelin metabolism, which is perturbed in CD, likely perturbing intestinal homeostasis^59^. We also find enriched pathogenic variants in genes within IBD-associated regions, though association tests lacked power to reach statistical significance, consistent with their low frequencies. Specifically, a pathogenic variant in *ARSA* (c.495_501del) implicated in metachromatic leukodystrophy shows a high enrichment in the UNRFC subset (AF=0.0019, biased corrected OR=88.6, one-sided Fisher exact test p-value=3.74 x 10^−11^). Similarly, although we do not find significant association in *AIRE*, a pathogenic variant in this gene (variant c.1616C>T implicated in autoimmune polyendocrinopathy-candidiasis-ectodermal dystrophy (APECED)^60^, shows an extremely high enrichment in the UNRFC cohort (AF=0.0028, biased-corrected OR=218.1, one-sided Fisher exact test p-value=2.5 x 10^−17^).

In addition to these single variant analyses, we performed burden tests on the sequencing data. These analyses found significant association to both the *NOD2* and *IL23R* genes (Table 2, Supplementary Table 17), consistent with previous findings that multiple variants in these genes are associated with IBD^61^. Two other genes had similarly significant burden test results as *NOD2*; these were the zinc finger protein *ZNF236* and the Citramalyl-CoA Lyase (*CLYBL*) genes (Table 2). The protein encoded by *ZNF236* is poorly characterized, although it is predicted to be involved in transcription or transcriptional regulation, with a preliminary link with autophagy^62^. In contrast, *CLYBL* is a well-characterized mitochondrial enzyme that is involved in the catabolism of itoconate, a metabolite of the TCA cycle with multiple immunomodulatory effects^63^.

**Table 2.**
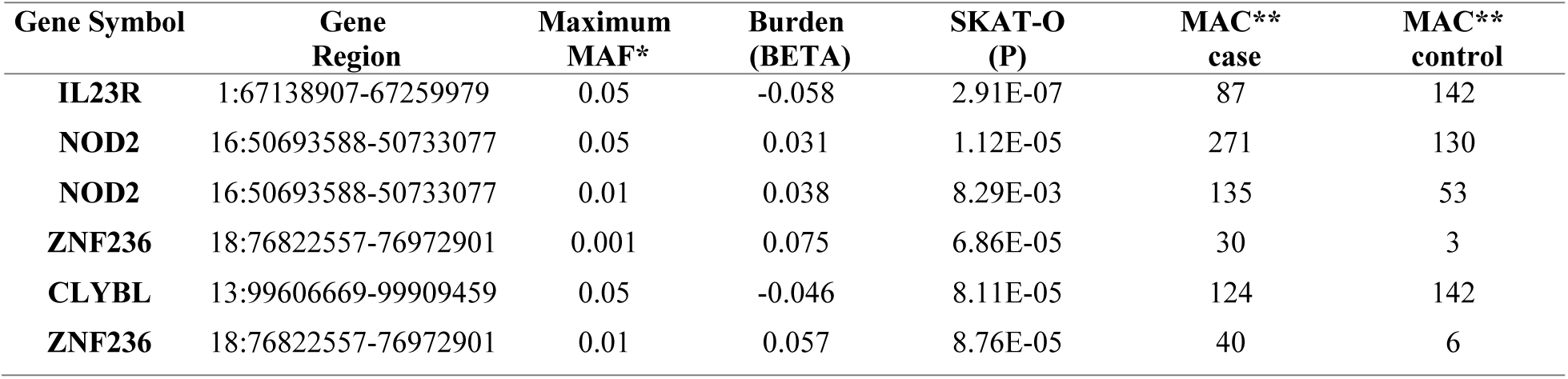
Summary of top burden test results.

| Gene Symbol | Gene Region | Maximum MAF* | Burden (BETA) | SKAT-O (P) | MAC** case | MAC** control |
| --- | --- | --- | --- | --- | --- | --- |
| **IL23R** | 1:67138907-67259979 | 0.05 | -0.058 | 2.91E-07 | 87 | 142 |
| **NOD2** | 16:50693588-50733077 | 0.05 | 0.031 | 1.12E-05 | 271 | 130 |
| **NOD2** | 16:50693588-50733077 | 0.01 | 0.038 | 8.29E-03 | 135 | 53 |
| **ZNF236** | 18:76822557-76972901 | 0.001 | 0.075 | 6.86E-05 | 30 | 3 |
| **CLYBL** | 13:99606669-99909459 | 0.05 | -0.046 | 8.11E-05 | 124 | 142 |
| **ZNF236** | 18:76822557-76972901 | 0.01 | 0.057 | 8.76E-05 | 40 | 6 |

## 5 Discussion

The French-Canadian population in Quebec has long been recognized as a prime example of a recent founder population with enrichment of certain Mendelian disease-causing variants^9–11^. A broader examination of coding variation in the French-Canadian population in Quebec, however, has long been limited to cohorts of a few hundred samples, or with low coverage sequencing, and in clinical settings with targeted sequencing technologies^12–15^. Here using exome sequencing data from 2,820 samples of French-Canadian ancestry, we provide a systematic analysis of coding variation in this founder population and its potential implications for Mendelian and common diseases.

Our study shows the enrichment of coding variants in French-Canadian exomes, with up to 45% having 4- to 100-fold enrichment in the UNRFC dataset as compared to the gnomAD NFE dataset. While we cannot compare this pattern directly to other founder populations without a joint variant call set, the observed enrichment pattern is similar to what was previously reported in other populations with recent bottleneck, including the Ashkenazi Jewish^44^ and the Finnish populations^64^. In our dataset, a large fraction of putatively deleterious variants was enriched more than 10-fold compared to NFE, precisely 31% of missense and 45% of predicted HC LoF variants. This is line with theory, where deleterious variation, usually maintained at lower frequencies by purifying selection, can increase to higher frequencies due to the founder effect, and this transient effect will be seen in populations where the bottleneck is relatively recent. This implies that if a deleterious variant is detected in such a population, then it has a greater probability of being enriched than in populations without a founder effect.

We show that previously reported founder variants are significantly enriched up to 76-fold in this French-Canadian subset of the Quebec IBD cohort compared to the gnomAD NFE reference sample. Our results also confirm the known regional concentration of some of these diseases, for instance the four diseases for which population screening is currently available in the SLSJ region^50^. We demonstrate that many other Mendelian disease-causing variants classified as pathogenic in ClinVar are enriched, yet to be described in the French-Canadian or Quebec context. Our study provides a comprehensive list of these variants and their annotations for future clinical and population-level follow-up. In the identified PLP variants at a MAF between 1/1000 to 1/100 with significant enrichment (one-sided Fisher exact test p-value < 0.05), there are 13 variants present in the list of 72 previously described founder variants, while 278 are not in the list and represent potential candidate founder variants. For example, we identified an enriched pathogenic variant in *FRAS1 (*p.Arg124Ter), implicated in Fraser syndrome 1 - a severe, autosomal recessive congenital malformation syndrome. This variant has not yet been described as founder variants in the French-Canadian population, and if confirmed, this has direct implications for genetic counselling. Our results also show a significant enrichment of the most common CHH-causing variant, *RMRP* n.70A>G. We also describe clinical data from probands of French-Canadian origin that corroborates the frequency and prevalence of CHH derived from French Canadian exome data. Although CHH is generally rare, it exhibits a high incidence in specific populations. The Old Order Amish population reports a prevalence of 1-2:1,000 (carrier frequency of 1:10), while in Finland, the prevalence is 1:23,000 (carrier frequency of 1:76)^52–54^. While the pathogenic variant n.70A>G is present in 100% of Old Order Amish, 92% of Finnish and 48% of non-Finnish individuals with CHH^52–54^, we show that it represents 80% of pathogenic variants in French Canadians. Among variants with PLP interpretations in ClinVar, we further show that only a small fraction is validated with two stars or more. This shows that a curation of pathogenic variants would be required to get a fuller picture of the landscape of known Mendelian disease-causing variants in the French-Canadian founder population.

Given these observations for known pathogenic variants for Mendelian diseases, we hypothesized that the French-Canadian population characteristics could also favor the discovery of some disease-causing alleles in common, complex diseases. To assess this possibility, we performed single variant association testing in the FC subset of the Quebec IBD cohort consisting of mother-father-affected child trios, affected individuals and unrelated controls. Consistent with previous large international studies, the top associations were linked to coding variants in the *NOD2* and *IL23R* genes^19,21,24^. For *IL23R*, of the three most associated coding variants from previous studies only one appeared to be enriched in this population (p.Val362Ile) whereas the other two were either underrepresented (p.Gly149Arg) or equivalent (p.Arg381Gln) to prevalence in NFE. For *NOD2*, all three of the top known coding variants (p.Arg702Trp; fs1007insC; p.Gly908Arg) were enriched 1.5-fold or greater in this population and significantly associated with CD. In a sequencing study of 30,000 patients with CD, a broader set of rare coding variants in *NOD2* associated with CD were identified^26^. Given the smaller size of the current cohort, not all of these rare variants were detected, but of the two that were, one was enriched in French Canadians (pSer431Leu) while the other was not (pVal793Met). These observations are consistent with a recent bottleneck in that not all coding variants are expected to pass through the bottleneck and that the degree of the enrichment will vary from one variant to another, although in general the degree of enrichment is less for common variants than rare variants. This suggests that although the allelic spectrum for disease-causing genes associated with common traits will be more limited in founder populations, the enrichment for certain alleles will favor their discovery in association studies. Indeed, single variant and burden tests in this French-Canadian cohort identified putative causal variants enriched in this population and associated with CD/IBD in genes not previously reported (e.g. *SLC35E3*, *ARSA*). Interestingly, all the candidate genes that we identified have functions that are linked with known pathophysiologic pathways of IBD such as autophagy, immune signaling, immunomodulation, and sphingomyelin metabolism (Supplementary Table 18). It will remain important to validate these novel findings, although in the short term functional studies will be needed given the lack of equivalently sized IBD replication cohort from this population.

All participants were recruited in the Province of Quebec, but as shown by our results, the Quebec IBD cohort comprised participants of diverse genetic ancestries. Thus, we used genetic similarity-based approach to define a French-Canadian subset and distinguish participants more genetically similar to the ERRQ reference sample, from participants more similar to the Europeans samples from the 1000 Genomes Project. While this definition of French Canadians excluded recently admixed individuals, it enabled us to capture known French-Canadian fine structure as the participants in the ERRQ were ascertained to represent a diversity of regional and ethno-cultural origins in Quebec^13^. Three main genetic clusters could be distinguished in our cohort, namely the northeastern Quebec cluster comprising the Saguenay−Lac-Saint-Jean regional population, the Acadian cluster from Eastern Quebec, and the Central Quebec cluster comprising metropolitan regions of Montreal and Quebec City. This was reflected in the distribution of carriers of ClinVar pathogenic variants in the PCA-based analyses. While this is not a systematic survey across the genome and the different populations from Quebec, our results highlight the regional differences in disease-causing founder variants and have practical implications for medical genetics services within this population. More broadly, this suggests that a finer understanding of regional differences of pathogenic variants could be beneficial, even in large heterogeneous populations.

In conclusion, even in well-characterized founder populations like the French Canadians that have enabled the mapping of many Mendelian disease genes, there remains untapped potential for genetic discovery of causal genes for rare diseases and complex disease risk factors. Large-scale sequencing studies across multiple founder populations will not only identify the individual variants enriched in each, but combining the evidence from different variants in the same genes or pathways across these studies will provide valuable information regarding the genetic architecture of disease across populations, as well as population-specific information for effective genetic counseling and public health needs.

## Data Availability

Sequence data used in this study have been made publicly available in dbGaP Study Accession number phs001642.v1.p1, Center for Common Disease Genomics (CCDG), Autoimmune: Inflammatory Bowel Disease (IBD) Exomes and Genomes (https://www.ncbi.nlm.nih.gov/projects/gap/cgi-bin/study.cgi?study_id=phs001642.v1.p1).

## 6 Consortia

### The Quebec IBD Genetics Consortium (QIGC)

Alain Bitton^1^, Guy Aumais^2^, Edmond-Jean Bernard^3^, Gabrielle Boucher^4^, Albert Cohen^5^, Colette Deslandres^6^, Gilles Jobin^2^, Raymond Lahaie^3^, Diane Langelier^7^, Pierre Paré^8^, Gary Wild^1^, John D. Rioux^4^

^1^ McGill University Health Centre, Montreal, Quebec, Canada; ^2^ Gastroentérologie, Hôpital Maisonneuve-Rosemont, Montréal, Quebec, Canada; ^3^ Centre Hospitalier de l’Université de Montréal, Montréal, Quebec, Canada; ^4^ Montreal Heart Institute and Université de Montréal, Montréal, Quebec, Canada; ^5^ Division of Gastroenterology, Jewish General Hospital, Montreal Quebec, Canada; ^6^ CHU-Sainte Justine, Montréal, Québec, Canada; ^7^ CHUS-Hôtel-Dieu, Sherbrooke, Quebec, Canada; ^8^ Hôpital Saint-Sacrement, Québec, Canada

### The iGenoMed Consortium

Robert Battat^1^, Alain Bitton^2^, Karine Tremblay^3^, Julie Thompson Legault^4^, Gabrielle Boucher^4^, Virginie Mercier^4^, Marie-Ève Rivard^4^, Pierre Poitras^1^, Justin Côté-Daigneault^1^, Katarzyna Orlicka^1^, Audrey Weber^1^, Benoît Panzini^1^, Edmond-Jean Bernard^1^, Érik Deslandres^1^, Louise D’Aoust^1^, Michel Boivin^1^, Michel Lemoyne^1^, Mickael Bouin^1^, Raymond Lahaie^1^, Raymond Leduc^1^, Simon Bouchard^1^, Alexia Monges^1^, Nancy Nadeau^1^, Stéphanie Côté-Daigneault^1^, Aimélie Bilodeau^1^, Louise Merly^1^, Karine Paupert^1^, Inès Ouchetati^1^, Bob Useni^1^, Marie-Pier Verpaelst^1^, Gary Wild^2^, Peter Lakatos^2^, Talat Bessissow^2^, Waqqas Afif^2^, Ernest Seidman^2^, Rita Kohen^2^, Kayleigh Morris^2^, Carolyne Lemieux^2^, Myriam Ouellette-Cabana^2^, Sakina Bennaghmouch^2^, Alban Michaud-Herbst^3^, Laurie Morin^3^, Jean-Maxime Picard^3^, Laurence Tessier^3^, Anne-Pascale Tremblay^3^, Joanie Bouchard^3^, John D. Rioux^4^

^1^ Centre Hospitalier de l’Université de Montréal, Montréal, Quebec, Canada; ^2^ McGill University Health Centre, Montreal, Quebec, Canada;^3^ CIUSSS Saguenay−Lac-Saint-Jean, Chicoutimi, Québec, Canada; ^4^ Montreal Heart Institute and Université de Montréal, Montréal, Quebec, Canada;

## 7 Data and Code Availability

Sequence data used in this study have been made publicly available in dbGaP Study Accession number phs001642.v1.p1, Center for Common Disease Genomics (CCDG), Autoimmune: Inflammatory Bowel Disease (IBD) Exomes and Genomes (https://www.ncbi.nlm.nih.gov/projects/gap/cgi-bin/study.cgi?study_id=phs001642.v1.p1). The analysis pipelines and custom scripts are available through version control repositories listed in the table below.

| Name | URL | Description |
| --- | --- | --- |
| vep_pipeline | https://github.com/CERC-Genomic-Medicine/vep_pipeline | A scalable Nextflow pipeline for variant annotation with Ensembl Variant Effect Predictor (VEP) |
| FC IBD paper | https://github.com/cbherer/FC_IBD_paper/ | Scripts to generate figures and results |

## 9 Acknowledgements

We thank all study participants for making this work possible. The authors would like to thank Julie Thompson Legault and Claire Le Moigne for their administrative support. This work was supported by the International IBD Genetics Consortium (IBDGC). Recruitment of study participants (patients and/or controls) was funded by the US National Institutes of Health grants no. DK062432 (JDR), the Fondation de l’Institut de cardiologie de Montréal (JDR), as well as a grant funded by Génome Québec, Genome Canada, the Government of Canada, and the Ministère de l’enseignement supérieur, de la recherche, de la science et de la technologie du Québec, the Canadian Institutes of Health Research (with contributions from the Institute of Infection and Immunity, the Institute of Genetics, and the Institute of Nutrition, Metabolism and Diabetes), Genome BC, and Crohn’s Colitis Canada via the 2012 Large-Scale Applied Research Project competition (grant # GPH-129341; JDR). Exome sequencing was funded in whole, or in part, by the US National Institutes of Health grants no. U54HG003067 (MJD) and no. 5UM1HG008895 (MJD) and The Leona M. & Harry B. Helmsley Charitable Trust grant no. 2015PG-IBD001 (MJD). We thank the Broad Institute Genomics Platform for genomic data generation efforts. CB is a Junior 1 Scholar from Les Fonds de la recherche Québec Santé (FRQS). JDR holds a Tier 1 Canada Research Chair (#230625). CL holds a Tier 1 Canada Research Chair (#2100108). VM holds a Canada Excellence Research Chair. This research was enabled in part by support provided by Digital Research Alliance of Canada (https://alliancecan.ca).

## 10 Author’s contributions

CB, JDR, VM, JH design and supervision of the project. CB, JCG, JP, GB, PG, DT sequencing data QC, statistical analyses and data interpretation. GG and DAB pathogenic variant curation. JDR, AB and Quebec IBD consortium participant recruitment, sample collection and management. CL and PC sharing of existing datasets. JDR, MD, CS, HH, sequencing data production and initial data processing. CB, JDR, JP, GB manuscript drafting. All authors: approval of the final version of the manuscript.

## 11 Web Resources

1000 Genomes Project, https://www.internationalgenomes.org

gnomAD, https://gnomad.broadinstitute.org/

PLINK 2.0, https://www.cog-genomics.org/plink/2.0/

FlashPCA2, https://github.com/gabraham/flashpca

Variant Effect Predictor (VEP), https://useast.ensembl.org/info/docs/tools/vep/index.html

LOFTEE, https://github.com/konradjk/loftee

