## Supplementary Material for "Frequency enrichment of coding variants in a French-Canadian founder population and its implication for inflammatory bowel diseases"

### Supplementary Figures

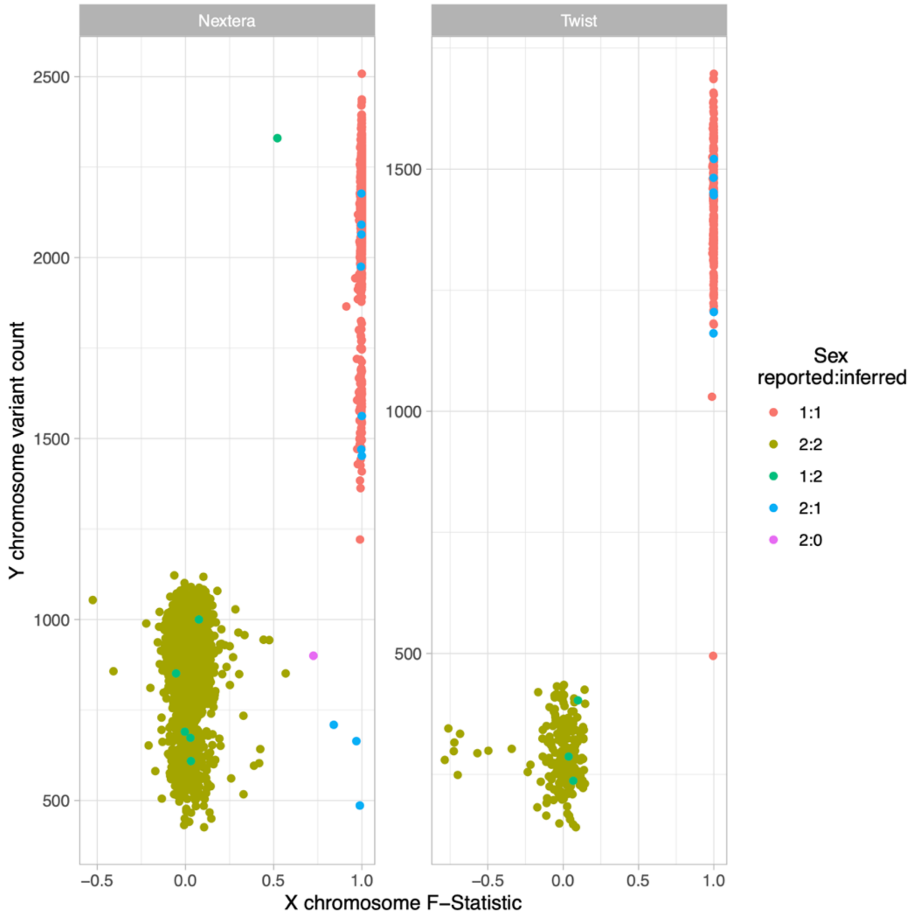


**Supplementary Figure 1. Genetic sex inference.** The X chromosome F-statistic was obtained with PLINK v1.9 using the --impute-sex 0.7 0.8 and --split-x options on a LD pruned dataset (--indep-pairwise 50 5 0.5) of common variants (MAF> 0.01) allowing for 10% missing rate. The number of non-missing Y chromosome variants were counted for each sample. Sex inference was performed separately for samples sequenced with Nextera and Twist captures (right and left panels, respectively). Each point represents one sample coloured according to sex reported in the metadata file (first number in the legend) versus sex inferred from genetic data (second number), where 1 refers to male, 2 to female and 0 to undetermined.

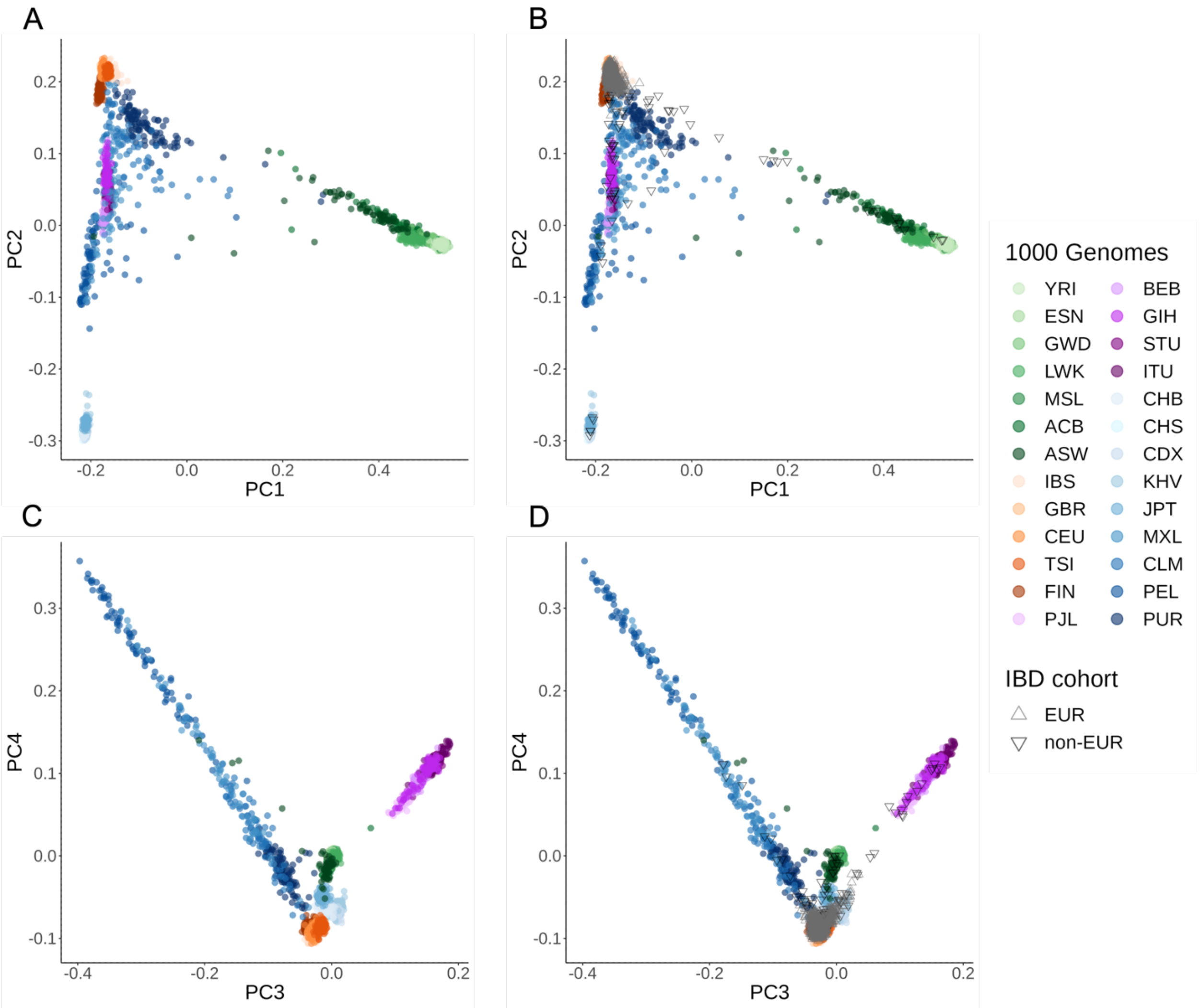


**Supplementary Figure 2. Principal component analysis (PCA) analysis of worldwide populations.** PCA of the 1000 Genomes and PCA projection of 3,015 samples from the Quebec IBD cohort. Upper panels (A-B) show PC1 vs PC2 and lower panels (C-D) PC3 vs PC4. The 1000 Genomes samples are represented by points colored according to 1000 Genomes superpopulation labels (AFR=green; EUR=orange; SAS=purple; AMR=blue). Sample abbreviations for the 1000 Genomes project are listed in Supplementary Table 3. Individuals from the Quebec IBD cohort (IBD cohort) are represented by up and down triangle symbols, respectively for individuals inferred most genetically similar to European (EUR) and non-European (non-EUR) samples from the 1000 Genomes project.

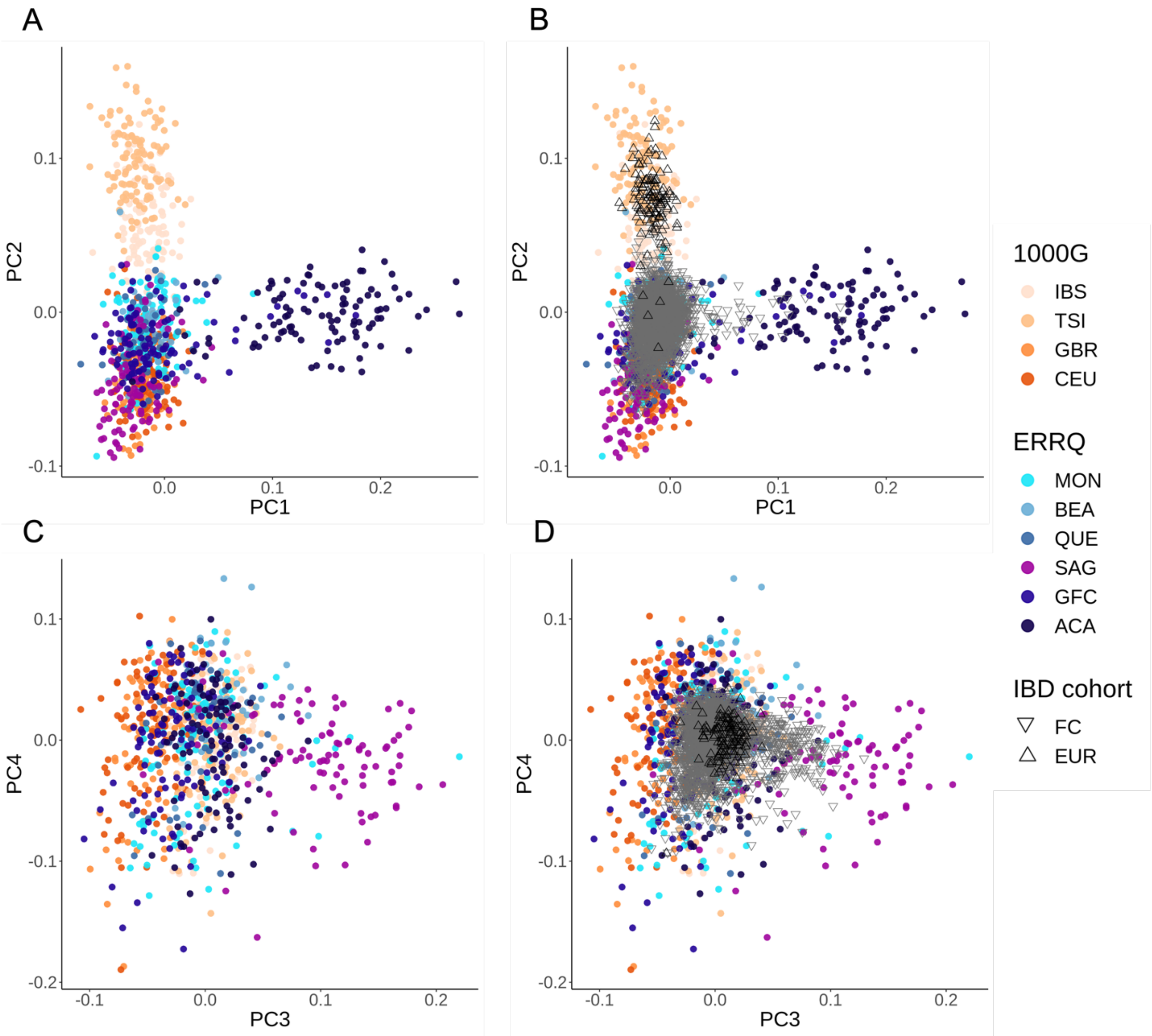


**Supplementary Figure 3. PCA of samples with European ancestries.** PCA of European population samples from the 1000 Genomes and Quebec samples from the ERRQ reference panel and PCA projection of IBD cohort samples. PCA is performed on a combined set of 4 European populations from the 1000 Genomes (excluding the Finnish sample) and a subset of the ERRQ. Upper panels show (A-B) PC1 vs PC2 and lower panels (C-D) PC3 vs PC4, with their respective proportion of variance explained (in %). Individuals from the IBD cohort are represented by down and up triangle symbols, respectively for individuals inferred most genetically similar to French Canadians (FC) from the ERRQ reference panel and to Europeans (EUR) population samples from the 1000 Genomes. Sample abbreviations for the 1000 Genomes project are listed in Supplementary Table 3, and those for the ERRQ in Supplementary Table 4.

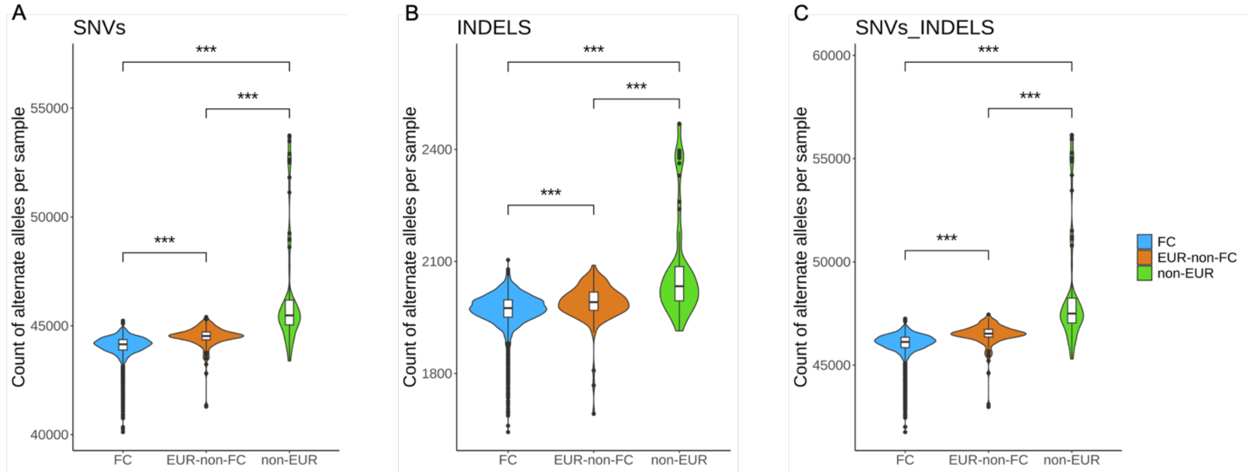


**Supplementary Figure 4. Distribution of variants in three subsets of the Quebec IBD cohort.** Violin plots represent the distribution of alternate alleles in three subsets of the Quebec IBD cohort, namely the FC subset, the EUR-non-FC and non-EUR subsets. Boxplots represent the median of the distribution and range between the first quartile (Q1, 25th percentile) and the third quartile (Q3, 75th percentile) and outliers are defined as datapoints outside of the smallest and largest values within the range defined as 1.5 times the IQR from the quartiles. Distributions were compared with a pairwise Wilcoxon test. All comparisons were significant (p-value < 0.001).

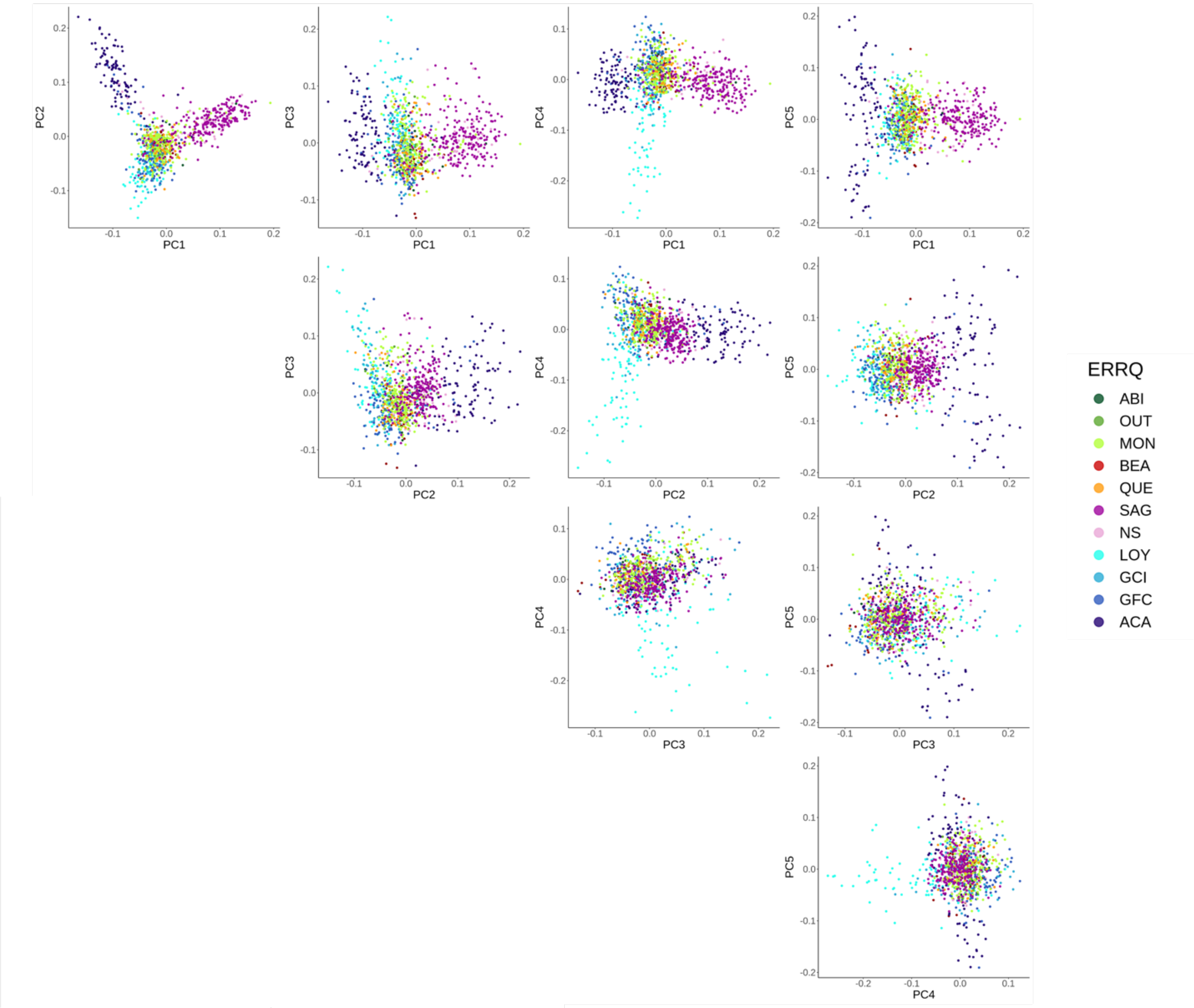


**Supplementary Figure 5. Fine-scale structure of French Canadians using the ERRQ reference panel.** PCA of the ERRQ reference panel. PC1 against PC5 are shown. Abbreviations: ABI: Abitibi-Témiscamingue; OUT: Outaouais; MON: Montreal; BEA: Beauce; QUE: Quebec City; SAG: Saguenay–Lac-Saint-Jean; NS: North-Shore; LOY: Gaspesia Loyalist; GCI: Gaspesia Channel Islanders; GFC: Gaspesia French Canadians; ACA: Gaspesia Acadians.

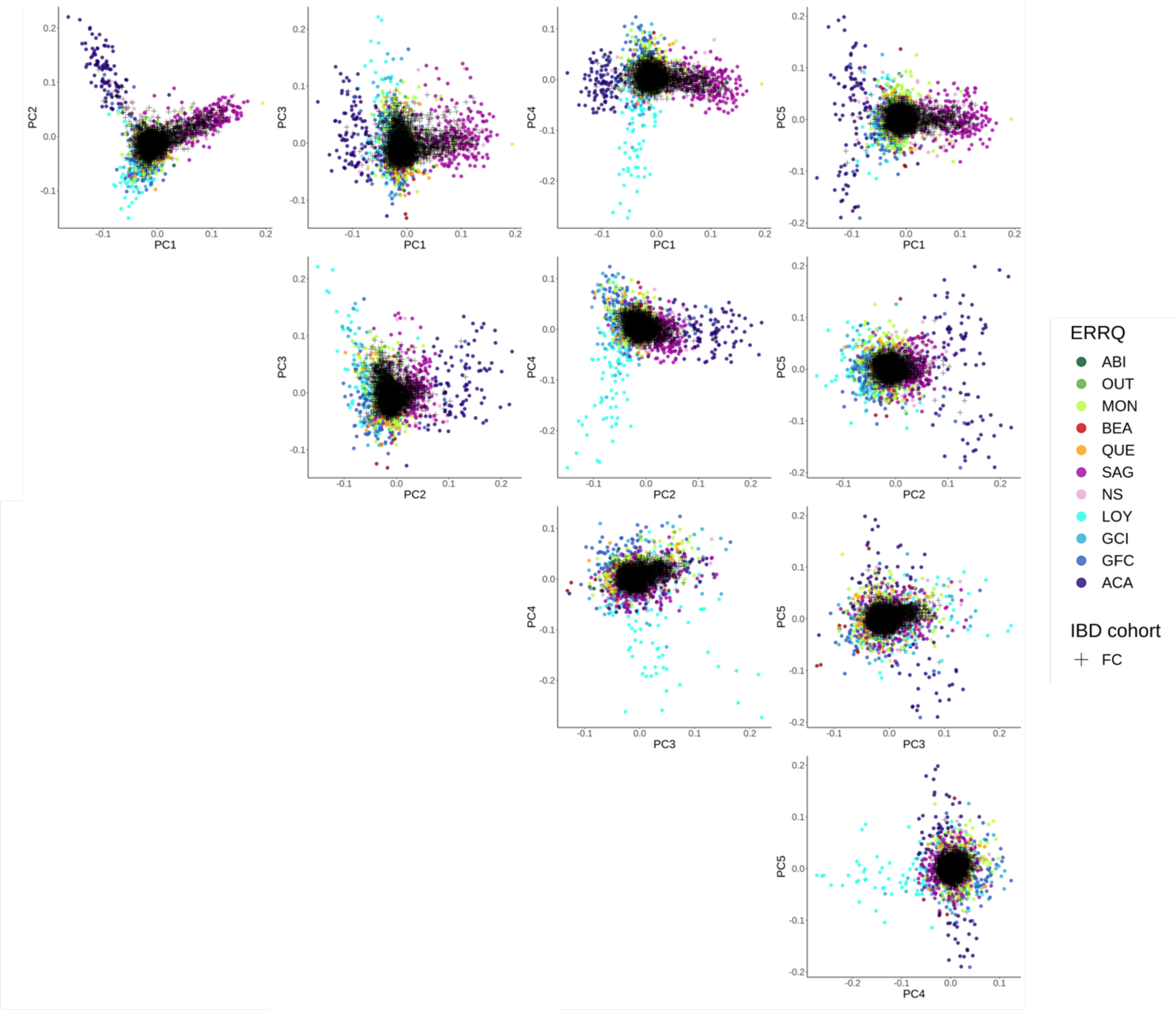


**Supplementary Figure 6. Fine-scale structure of French Canadians using the ERRQ reference panel and projection of the FC subset.** PCA of the ERRQ reference panel and projection of the FCIBD subset. PC1 against PC5 are shown. Abbreviations: ABI: Abitibi-Témiscamingue; OUT: Outaouais; MON: Montreal; BEA: Beauce; QUE: Quebec City; SAG: Saguenay–Lac-Saint-Jean; NS: North-Shore; LOY: Gaspesia Loyalist; GCI: Gaspesia Channel Islanders; GFC: Gaspesia French Canadians; ACA: Gaspesia Acadians, FC: French-Canadians

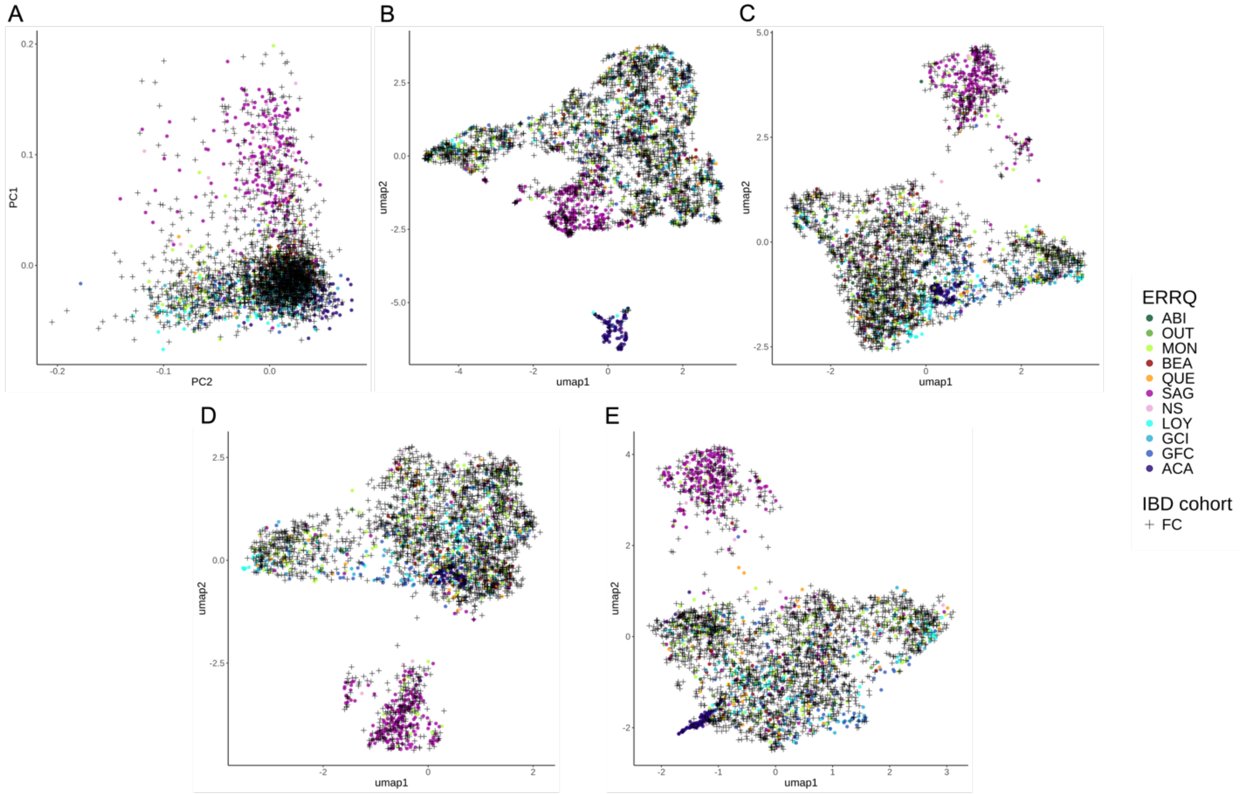


**Supplementary Figure 7**. **Uniform Manifold Approximation and Projection (UMAP) of the ERRQ and UNRFC samples.** (A) Principal component analysis representing PC1 and PC2. B) UMAP of the top 5 PCs. (C) UMAP of the top 10 PCs. D) UMAP of the top 15 PCs. E) UMAP of the top 20 PCs. Abbreviations: ABI: Abitibi-Témiscamingue; OUT: Outaouais; MON: Montreal; BEA: Beauce; QUE: Quebec City; SAG: Saguenay–Lac-Saint-Jean; NS: North-Shore; LOY: Gaspesia Loyalist; GCI: Gaspesia Channel Islanders; GFC: Gaspesia French Canadians; ACA: Gaspesia Acadians, FC: French-Canadians

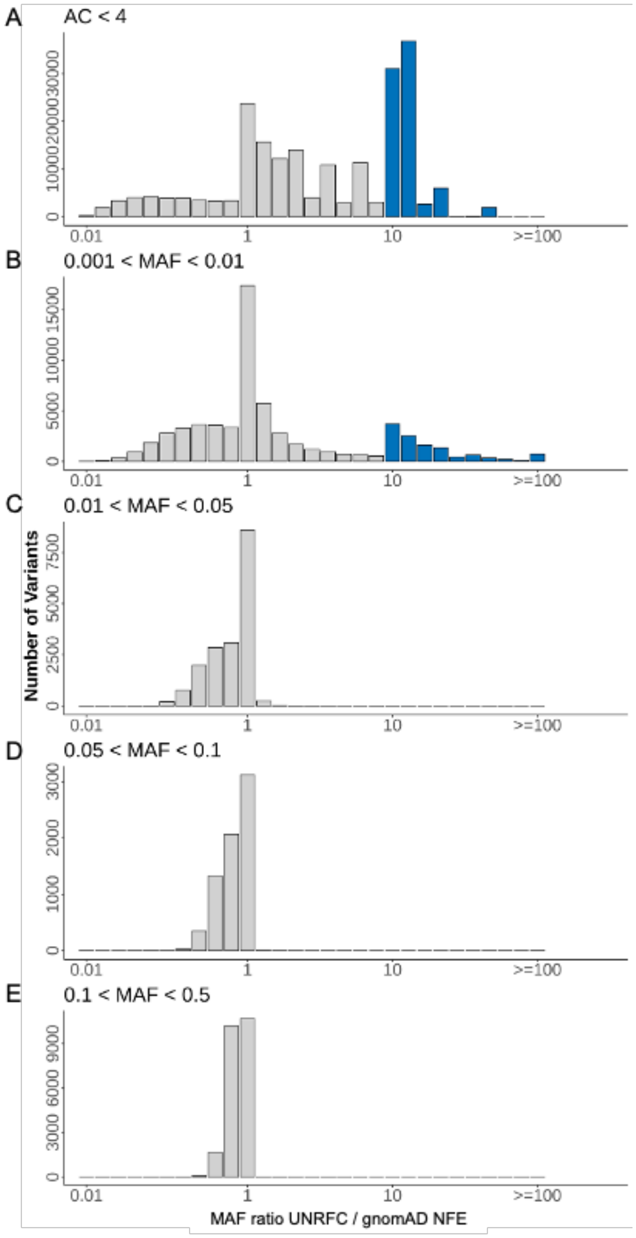


**Supplementary Figure 8. Enrichment of protein-coding variants per frequency categories.** Histograms show the distribution of variants as a function of their minor allele frequency (MAF) ratio in the UNRFC sample relative to gnomAD NFE exomes. Variants with MAF ratio equal or above 10-fold enrichment in the UNRFC are highlighted in blue. Frequency enrichment histograms are shown for variants in different MAF categories in the UNRFC subset, specifically (A) allele count (AC) < 4 (MAF = 0.001), (B) 0.001 < MAF < 0.01, (C) 0.01 < MAF < 0.05, (D) 0.05 < MAF < 0.1, (E) 0.1 < MAF < 0.5.

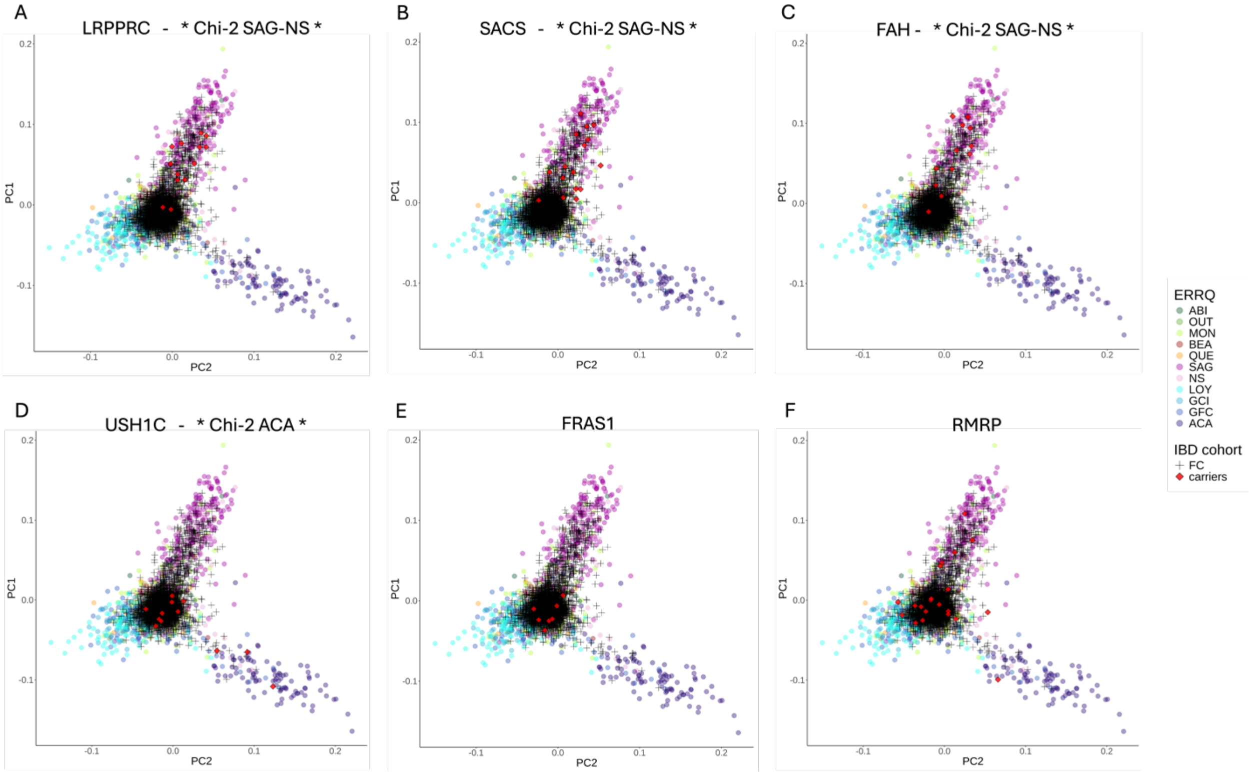


**Supplementary Figure 9. PCA plots highlight carriers of PLP variants in the UNRFC subset.** The PCA was performed on the ERRQ reference panel, and the exomes from the UNRFC subset were then projected on this PCA. Carriers of PLP variants are highlighted in red. Abbreviations: ABI: Abitibi-Témiscamingue; OUT: Outaouais; MON: Montreal; BEA: Beauce; QUE: Quebec City; SAG: Saguenay–Lac-Saint-Jean; NS: North-Shore; LOY: Gaspesia Loyalist; GCI: Gaspesia Channel Islanders; GFC: Gaspesia French Canadians; ACA: Gaspesia Acadians, FC: French-Canadians.

### Supplementary Tables

**Supplementary Table 1.** Summary of the Quebec IBD cohort by study cohort before and after quality control (QC). Study cohorts are described in Materials and Methods and include IBDGC-Montreal, iGenomed-MTT, Genome Quebec-GENIZON Case-Control (GC_CC), Genome Quebec-GENIZON Trios (GC_Trios).

| Study cohort | Before QC | After QC |
|---|---|---|
| IBDGC-Montreal | 995 | 931 |
| IGenomed-MTT | 186 | 169 |
| GC_CC | 872 | 869 |
| GC_Trios | 1,049 | 1,046 |
| Total | 3,102 | 3,015 |

**Supplementary Table 2.** Summary of quality control steps and filtering.

| | Number of samples | Number of variants | Genotyping rate | Number of sites removed |
|---|---|---|---|---|
| **Initial variant callset** | **3,102** | **4,435,411** | **0.6784** | |
| **Genotype-level filters** | | | | |
| Genotype quality > 20 | 3,102 | 4,435,411 | 0.5890 | - |
| PASS | 3,102 | 3,826,429 | 0.5816 | 608,982 |
| Allele balance ratio | 3,102 | 3,826,429 | 0.5793 | - |
| Normalize InDels | 3,102 | 3,826,429 | 0.5793 | - |
| **Individual-level filters** | | | | |
| No Duplicated Samples | 3,074 | 3,826,429 | 0.5794 | - |
| No Sex Mismatch, chimera, contamination | 3,015 | 3,826,429 | 0.5805 | - |
| **Genotype-/variant- level filters** | | | | |
| **Autosomes** | **3,015** | **3,729,594** | **0.5806** | |
| DP > 10 | 3,015 | 3,729,594 | 0.5466 | - |
| Mendelian errors set as missing | 3,015 | 3,729,594 | 0.5466 | - |
| Missing < 5% | 3,015 | 1,258,779 | 0.9928 | 2,470,815 |
| Hardy Weinberg p-value < 1 x 10-6 | 3,015 | 1,258,415 | 0.9928 | 364 |
| Matching exome kits with 50bp flanks | 3,015 | 848,032 | 0.9956 | 410,383 |
| **Total after quality control filters** | 3,015 | 848,032 | 0.9956 | - |

**Supplementary Table 3.** 1000 Genomes cohort populations description. The superpopulation labels stand for: AFR = Africa, AMR = America, EAS = East Asia, EUR = Europe, SAS = South Asia.

| Sample abbreviation | Superpopulation | Description |
|---|---|---|
| ACB | AFR | African Caribbean in Barbados |
| ASW | AFR | African Ancestry in Southwest US |
| BEB | SAS | Bengali in Bangladesh |
| CDX | EAS | Chinese Dai in Xishuangbanna, China |
| CEU | EUR | Utah residents with Northern and Western European ancestry |
| CHB | EAS | Han Chinese in Bejing, China |
| CHS | EAS | Southern Han Chinese, China |
| CLM | AMR | Colombian in Medellin, Colombia |
| ESN | AFR | Esan in Nigeria |
| FIN | EUR | Finnish in Finland |
| GBR | EUR | British in England and Scotland |
| GIH | SAS | Gujarati Indian in Houston,TX |
| GWD | AFR | Gambian in Western Division, The Gambia |
| IBS | EUR | Iberian populations in Spain |
| ITU | SAS | Indian Telugu in the UK |
| JPT | EAS | Japanese in Tokyo, Japan |
| KHV | EAS | Kinh in Ho Chi Minh City, Vietnam |
| LWK | AFR | Luhya in Webuye, Kenya |
| MSL | AFR | Mende in Sierra Leone |
| MXL | AMR | Mexican Ancestry in Los Angeles, California |
| PEL | AMR | Peruvian in Lima, Peru |
| PJL | SAS | Punjabi in Lahore,Pakistan |
| PUR | AMR | Puerto Rican in Puerto Rico |
| STU | SAS | Sri Lankan Tamil in the UK |
| TSI | EUR | Toscani in Italy |
| YRI | AFR | Yoruba in Ibadan, Nigeria |

**Supplementary Table 4.** ERRQ cohort regional and ethno-cultural groups description.

| Abbreviation | Regional or ethno-cultural population | Geographical Region in Quebec | Number of samples |
|---|---|---|---|
| ABI | Abitibi | North-West | 24 |
| OUT | Outaouais | West | 17 |
| MON | Montreal | Central | 215 |
| BEA | Beauce | Central | 44 |
| QUE | Quebec | Central | 53 |
| SAG | Saguenay–Lac-Saint-Jean | North-East | 232 |
| NS | North-Shore | North-East | 71 |
| LOY | Gaspesia Loyalist | East | 94 |
| GCI | Gaspesia Channel-Islanders | East | 91 |
| GFC | Gaspesia French-Canadian | East | 106 |
| ACA | Gaspesia Acadia | East | 101 |

**Supplementary Table 5.** Detailed description of IBD phenotypes in the FC subset.

| | Case/Control | | | Trios |
|---|---|---|---|---|
| | **IBD*** | **CD** | **UC** | **CD index** |
| Total | 1016 | 768 | 228 | 318 |
| **Sex** | | | | |
| Female | 582 | 454 | 117 | 205 |
| Male | 434 | 314 | 111 | 113 |
| **Age at diagnosis** | | | | |
| Median [Q1-Q3] | 27 [19-40] | 27 [19-40] | 28 [21-40] | 21[17-26] |
| Missing | 38 | 32 | 6 | 5 |
| **Disease Location CD** | | | | |
| Ileal (L1) | - | 219 | - | 87 |
| Colorectal (L2) | - | 192 | - | 76 |
| Ileocolonic (L3) | - | 333 | - | 155 |
| Missing | - | 24 | - | 0 |
| **Disease Behavior CD** | | | | |
| Inflammatory (B1) | - | 378 | - | 142 |
| Stricturing (B2) | - | 145 | - | 57 |
| Penetrating (B3) | - | 222 | - | 119 |
| Missing | - | 23 | - | 0 |
| **Perianal CD** | | | | |
| Yes | - | 102 | - | 0 |
| No | - | 293 | - | 0 |
| Missing | - | 373 | - | 318 |
| **Disease Extent UC** | | | | |
| Proctitis (E1) | - | - | 27 | - |
| Left-Sided (E2) | - | - | 104 | - |
| Extensive (E3) | - | - | 89 | - |
| Missing | - | - | 9 | - |

*Indeterminate Colitis (n=20) is included in the IBD subset with CD and UC cases

**Supplementary Table 6.** Variant summary for the Quebec IBD cohort. Number of variants, SNVs and InDels after quality controls. Novel variants are those not in dbSNP.

| Variants | All (N) | All (% singletons) | Novel (N) | Novel (% singletons) | In dbSNP (N) | In dbSNP (% singletons) |
|---|---|---|---|---|---|---|
| All | 848,032 | 40.96 | 52,994 | 58.98 | 795,038 | 39.76 |
| SNV | 804,034 | 41.23 | 46,082 | 62.21 | 757,952 | 39.96 |
| InDel | 43,998 | 36.00 | 6,912 | 37.44 | 37,086 | 35.73 |

**Supplementary Table 7.** Summary of exome data by subset of the Quebec IBD cohort. The FC subset contains individuals inferred most genetically similar to FC reference samples. These samples were included in single-point association test and burden test. The UNRFC subset contains individuals inferred most genetically similar to FC reference samples, unrelated up to the third degree. The UNRFC subset was used in enrichment analyses. The FC controls subset contains control-only individuals inferred most genetically similar to FC reference samples. This subset was used to compute enrichment statistics for variants identified in association tests. The EUR-non-FC subsets and non-EUR subsets were used to describe the distribution of coding variants per exomes.

| | Number of samples | Number of variants | | |
|---|---|---|---|---|
| | | All | SNVs | InDels |
| All | 3,015 | 848,032 | 804,034 | 43,998 |
| FC | 2,820 | 653,574 | 623,119 | 30,455 |
| UNRFC | 2,323 | 640,073 | 610,326 | 29,747 |
| FC controls | 760 | 369,924 | 352,747 | 17,177 |
| EUR-non-FC | 127 | 203,128 | 193,870 | 9,258 |
| Non-EUR | 68 | 227,573 | 217,357 | 10,216 |

**Supplementary Table 8.** Descriptive statistics of the Quebec IBD cohort, the FC and the UNRFC subsets.

| **Demographic and clinical characteristics** | **All** | **FC** | **UNRFC** |
|---|---|---|---|
| Number of participants | 3,015 | 2,820 | 2,323 |
| Number of women (%) | 1,726 (57.25) | 1,615 (57.30) | 1,321 (56.87) |
| Number of IBD patients | 1,507 | 1,364 | 1,002 |
| Number of parents | 699 | 696 | 637 |
| Number of controls | 809 | 760 | 684 |
| **IBD phenotypes of patients** | | | |
| Crohn's disease | 1,207 | 1,116 | 769 |
| Ulcerative colitis | 275 | 228 | 214 |
| Indeterminate colitis | 25 | 20 | 19 |

**Supplementary Table 9.** Number of variants per exome in the FC, non-EUR, and EUR-non-FC subsets. The mean number of variants per exome is shown, as well as standard deviation (s.d.), variance, median and 5th and 95th percentile of the distribution.

| Variant type | Population | mean | s.d. | variance | median | 5th perc | 95th perc |
|---|---|---|---|---|---|---|---|
| SNVs + InDels | FC | 46,021.86 | 577.09 | 333,034.55 | 46,126.00 | 44,984.95 | 46,649.15 |
| | non-EUR | 48,536.69 | 2,858.40 | 8,170,426.01 | 47,499.00 | 46,067.00 | 55,189.95 |
| | EUR-non-FC | 46,450.68 | 601.31 | 361,575.68 | 46,523.00 | 45,549.40 | 47,121.50 |
| | ALL | 46,096.64 | 808.13 | 653,070.10 | 46,146.00 | 45,037.10 | 46,765.60 |
| SNVs | FC | 44,053.21 | 539.56 | 291,128.73 | 44,149.00 | 43,087.95 | 44,653.05 |
| | non-EUR | 46,456.59 | 2,721.68 | 7,407,523.29 | 45,477.00 | 44,109.40 | 52,805.30 |
| | EUR-non-FC | 44,462.22 | 560.82 | 314,515.25 | 44,529.00 | 43,564.90 | 45,105.70 |
| | ALL | 44,124.65 | 763.13 | 582,371.61 | 44,171.00 | 43,135.70 | 44,766.30 |
| InDels | FC | 1,968.64 | 49.55 | 2,454.84 | 1,975.00 | 1,882.00 | 2,030.00 |
| | non-EUR | 2,080.10 | 140.42 | 1,9716.66 | 2,033.50 | 1,949.40 | 2,388.55 |
| | EUR-non-FC | 1,988.46 | 50.99 | 2,600.35 | 1,991.00 | 1,936.20 | 2,054.00 |
| | ALL | 1,971.99 | 55.93 | 3,128.63 | 1,976.00 | 1,885.00 | 2,036.00 |

**Supplementary Table 10.** Variant summary of coding variants by consequence in the UNRFC subset.

| Variants | All (N) | All (% singletons) | Novel (N) | Novel (% singletons) | In dbSNP (N) | In dbSNP (% singletons) |
|---|---|---|---|---|---|---|
| All | 398,626 | 45.7 | 17,759 | 90.8 | 380,867 | 43.6 |
| Synonymous | 138,836 | 40.9 | 4,325 | 90.4 | 134,511 | 39.3 |
| Non-synonymous | 246,067 | 48 | 11,848 | 91 | 234,219 | 45.9 |
| Splice | 3,017 | 56.3 | 346 | 91.3 | 2,671 | 51.8 |
| Frameshift | 6,147 | 56.8 | 927 | 90.6 | 5,220 | 50.8 |
| Inframe | 4,526 | 46 | 309 | 88.3 | 4,217 | 42.9 |

**Supplementary Tables in Excel File:** SUPP_Data1_PLP_variants.xlsx

**Supplementary Table 11.** Previously described French Canadian founder variants. Curated list of known pathogenic variants for Mendelian diseases previously described with founder effect in French Canadians.

**Supplementary Table 12.** ClinVar PLP variants. Variant annotations for ClinVar PLP variants in the UNRFC unrelated subset.

**Supplementary Table 13.** ClinVar PLP variants in the *RMRP* gene in the UNRFC subset.

| Variant (hg38) dbSNP | HGVSc | Consequence | ClinVar ID | ClinVar significance | AC | AN | NS | Carrier Rate |
|---|---|---|---|---|---|---|---|---|
| 9:35657801:G:A rs1021056174 | ENST00000363046.1 n.217C>T | Noncoding transcript exon variant | 2445847 | Variant of unknown significance - 1 star | 1 | 4646 | 2323 | 1/2323 |
| 9:35657872:C:T rs753874439 | ENST00000363046.1 n.146G>A | Noncoding transcript exon variant | 379208 | Pathogenic - 2 stars | 1 | 4646 | 2323 | 1/2323 |
| 9:35657948:T:C rs199476103 | ENST00000363046.1 n.70A>G | Noncoding transcript exon variant | 14208 | Pathogenic/Likely Pathogenic 2 stars | 12 | 4646 | 2323 | 1/194 |
| 9:35658014:G:C rs772443941 | ENST00000363046.1 n.4C>G | Noncoding transcript exon variant | 552477 | Pathogenic/Likely Pathogenic 2 stars | 1 | 4624 | 2312 | 1/2312 |

**Supplementary Table 14.** Pathogenic variants observed in RMRP in 5 probands diagnosed with CHH in a tertiary care center.

| VarID (hg38) | Preferred name | Consequence | ClinVar ID | ClinVar significance | AC | AN | AC (FC) | AN (FC) |
|---|---|---|---|---|---|---|---|---|
| 9:35657948:T:C rs199476103 | NR_003051.4(RMRP) n.72A>G | Noncoding transcript variant | 14208 | Pathogenic/Likely Pathogenic - 2 stars | 6 | 10 | 6 | 8 |
| 9:35657807:G:C rs192060920 | NR_003051.4(RMRP) n.213C>G | Noncoding transcript variant | 632970 | Pathogenic/Likely Pathogenic - 2 stars | 2 | 10 | 0 | 8 |
| 9:35657804:T:A - | NC_000009.12 g.35657804T>A | Noncoding transcript variant | 2078709 | Variant of unknown significance - 1 star | 1 | 10 | 1 | 8 |
| 9:35657825:C:T rs761398394 | NR_003051.4(RMRP) n.195G>A | Noncoding transcript variant | 928882 | Pathogenic/Likely Pathogenic - 2 stars | 1 | 10 | 1 | 8 |

**Supplementary Tables in Excel Files : SUPP_Data2_FC_IBD_Association.xlsx**

**Supplementary Table 15.** Association and enrichment results in known causal variants.
**Supplementary Table 16.** Novel association results.
**Supplementary Table 17:** SKAT-O Burden test results for the top 20 genes for non-synonymous and LoF variants.
**Supplementary Table 18.** Summary of key genes and function.
